# Nociceptive stimulation suppresses spinal beta oscillations in awake human epidural recordings

**DOI:** 10.64898/2026.04.21.26351212

**Authors:** Rahul S. Shah, Aditya M. Nair, Meredith Macdonald, Michael G. Hart, Erlick A. C. Pereira

**Author notes:** Correspondence to: Rahul S. Shah, Full Address: Department of Psychology & Neuroscience, City St George’s, University of London, Cranmer Terrace, London, SW17 0RE, UK.

## Abstract

**Introduction:** Nociceptive processing in the human spinal cord remains difficult to study directly, and its oscillatory dynamics are poorly understood. Oscillatory activity in the beta band (13–35 Hz) is of particular interest, as beta rhythms are widely associated with sensorimotor network state and transient desynchronisation following salient input.

**Objectives:** Identify the effects of nociceptive laser thermal stimuli on beta-band oscillatory activity in epidural spinal field potentials.

**Methods:** We recorded epidural spinal field potentials during noxious thermal laser stimulation of the unaffected foot using externalised thoracic spinal cord stimulation electrodes in two subjects with neuropathic pain (persistent spinal pain syndrome type 2).

**Results:** Time–frequency analysis combined with cluster-based permutation testing revealed reproducible suppression of spinal beta oscillations (13–35 Hz) following nociceptive stimulation. Beta suppression was spatially organised along contiguous rostro-caudal bipolar channels and most prominent within the first 0–200 ms after stimulation. Both low- (13-20 Hz) and high-beta (20-35 Hz) sub-bands contributed to early effects, with low-beta suppression predominating rostrally and while high-beta suppression was more ubiquitous. Inter-trial coherence increases were modest and not consistently aligned with early suppression, suggesting induced desynchronisation rather than a dominant phase-reset response.

**Conclusion:** Nociceptive input produces early, spatially organised modulation of spinal beta oscillation dynamics in awake subjects.

## Introduction

Spinal epidural field recordings in humans originated in early intraoperative neurophysiology with mid-20th century studies demonstrating that epidural or subdural electrodes could capture stimulus-evoked population activity from dorsal columns and roots, providing insight into spinal conduction and segmental organisation [14,24,25]. These observations underpinned the subsequent development of intraoperative spinal cord monitoring [26]. In parallel, the introduction of spinal cord stimulation (SCS) for pain created a chronic interface with the epidural space, enabling repeated recordings of evoked compound action potentials (ECAPs) and slower field responses from dorsal column and segmental generators [7,20]). More recently, advances in implantable hardware and signal processing have renewed interest in epidural spinal field potentials (eSFP) as a window into human spinal network dynamics in awake subjects [4,29]. While current research in SCS for motor rehabilitation primarily infers spinal activity indirectly through peripheral outputs [1,13], it is unclear whether eSFPs can capture rapid physiological oscillatory network dynamics analogous to those observed in cortical and subcortical systems, where rhythmic activity reflects coordinated network state rather than purely time-locked conduction. Unlike most intracranial recordings, epidural spinal electrodes are separated from neural tissue by dura and cerebrospinal fluid, leading to attenuation and spatial blurring of signals. Posture-dependent cord–electrode coupling, paraspinal muscle activity, and cardiorespiratory motion further complicate signal interpretation. Consequently, most epidural signal analysis has focused on ECAPs, which are large and time-locked [20].

Across cortical and subcortical systems, beta oscillations (13-35 Hz) are widely associated with maintenance of network state, sensorimotor integration, and top–down modulation, and are thought to reflect coordinated activity across distributed neuronal populations [5,11]. If similar dynamics exist within the spinal cord, beta band activity could provide a tractable marker of spinal network engagement that is detectable at the eSFP level despite the spatial and biophysical constraints of epidural recording. While spinal beta activity has been reported averaged over long time periods [4,29], direct evidence for stimulus-evoked beta modulation within the human spinal cord is absent.

Laser-evoked potentials (LEPs) provide a useful framework for addressing this gap. Cutaneous laser stimulation selectively activates nociceptive Aδ and C fibres without direct electrical spread to deeper tissues and produces a temporally precise afferent volley. Cortical LEPs are well characterised and include both lateralised components reflecting sensory-discriminative processing and medial potentials associated with salience and affective processing [8,27]. Importantly, the sharp temporal structure of LEPs enables separation of stimulus-locked responses from ongoing activity through averaging and time– frequency analysis [21]. Compared with mechanical, ramp thermal, or tonic pain paradigms, which generate temporally diffuse responses, laser stimulation provides a brief and physiologically specific input that is well suited for interrogating eSFP responses in the presence of epidural noise and state-dependent variability. This temporal precision makes laser stimulation particularly suitable for probing beta band dynamics, which may manifest as transient suppression or reorganisation rather than large evoked-potentials.

We recorded eSFP via implanted thoracic spinal cord stimulation electrodes during laser thermal stimulation in two subjects with neuropathic pain. By combining rostro-caudally resolved bipolar recordings with cluster-based permutation analysis, we sought to determine whether nociceptive input evokes reproducible beta-band modulation within human spinal networks.

## Methods

### Participants and surgery

The study was approved by the NHS Health Research Authority (IRAS 294531) and subjects gave informed written consent before the recording. SCS trial electrodes were implanted under general anaesthesia using fluoroscopic guidance. The implanted trial leads were directly externalised through the skin of the back, secured with silk sutures, and connected to an external stimulator. Subjects were then programmed by the device company representatives and discharged the following day with multiple programmes to test at home.

Both subjects were contacted prior to their end of trial visit and agreed to participate in the study so were instructed to turn off their stimulator >24h prior to coming to hospital to allow stimulation washout [16]. Subject characteristics are outlined in **Table 1**.

**Table 1.** Subject characteristics. PSPS, persistent spinal pain syndrome.

| Subject ID | Age range (years) | Sex | Diagnosis | Pain location | Pain Medications | Trial Leads | Response to SCS Trial | Best Program Leads | Best Program Pattern |
| --- | --- | --- | --- | --- | --- | --- | --- | --- | --- |
| 1<br>(crnp04) | 55-59 | M | PSPS Type II<br>Previous lumbar discectomy | Back and left leg. | Gabapentin 3.6g/d<br>Amitriptyline 75mg/d<br>Buprenorphine patch 15mcg/h | Single Saluda 12 ch with tip of lead bottom of T7 | 65% pain reduction | Top third of lead | ECAP-closed loop |
| 2<br>(crnp05) | 40-44 | F | PSPS Type II<br>Previous lumbar discectomy, fusion surgery and revision fusion surgery. | Back and left leg | Tapentadol 100mg qds<br>Codeine 60mg/ Paracetamol 1g mg qds<br>Nortriptyline 50mg<br>Lidocaine patches | Two Nevro 8 ch with leads tips at top of T8 and T9 | 65% pain reduction | Left (rostral) lead only | Cascade 10kHz |

### Laser-thermal stimuli recordings and analysis

eSFP recordings [29] and thermal laser stimulation [3,9] are described in Supplementary Data.

### Time-Frequency Analysis

Prior to analysis electrodes were classified anatomically to allow relative position of electrodes on adjacent midline leads to be localised in the same parameter space in addition to their known relative position within a SCS lead. After removal of trials with significant artefact, 24 trials in Subject 1 and 27 in Subject 2 were included for analysis. Two bipolar referencing schemes were analysed to balance spatial specificity against sensitivity to distributed field activity in eSFP recordings. Bipolar channels formed from every other contact increase inter-electrode distance and preferentially capture mesoscale field potentials, enhancing sensitivity to spatially extended or synchronised activity but at the cost of increased susceptibility to volume conduction. In contrast, adjacent-contact bipolar channels act as a spatial high-pass filter, reducing common-mode signals and volume-conducted contributions while improving spatial specificity, albeit with reduced sensitivity to low-spatial-frequency fields. Analysing both montages allowed us to assess whether observed oscillatory effects reflected focal neural generators or broader distributed fields, and to test the robustness of key findings to reference choices. Effects that persist across both montages are unlikely to arise from reference-dependent artefacts or diffuse volume conduction, whereas effects that attenuate or fragment with adjacent bipolar referencing likely reflect more spatially extended or lower-frequency field phenomena.

Time-frequency representations (TFRs) were calculated for each spinal bipolar channel and Cz-Ref channel using Fieldtrip function ‘ft_freqanalysis’ with multitaper spectral decomposition. Different spectral estimation approaches were applied to three main frequency bands to optimize time-frequency resolution:

- *Low Frequency Range (5-30 Hz*): Hanning tapers with adaptive frequency-dependent time windows (5 cycles per frequency) across 0.5 Hz steps, with time points sampled at 10 ms intervals. Data were zero-padded to 20 seconds to reduce boundary effects
- *Mid Frequency Range (20-100 Hz):* Discrete prolate spheroidal sequence (DPSS) multitapers with fixed time windows (0.5s) and moderate frequency smoothing (5 Hz) with time points sampled at 10ms intervals.
- *High Frequency Range (100-500 Hz):* DPSS multitapers with frequency-adaptive time windows (minimum 0.1s) and broader spectral smoothing (30 Hz) across 5Hz steps with time points sampled at 10ms intervals to capture HFO dynamics.

For Subject 1 (single Saluda 12 channel lead, but only top 11 channels recorded), TFRs were computed separately for adjacent-contact (10 channels) and every-other-contact (9 channels) bipolar montages. For Subject 2 (dual Nevro 8 channel leads), TFRs were computed separately for each lead and each montage, yielding two sets adjacent-contact (7 channels) and every-other-contact (6 channels) bipolar montages. The overall span of all leads from the centre of the top to bottom contact was ∼80mm. As such, for subject 1 the rostro-caudal span assessed was ∼80mm while for subject 2 it was ∼120mm due to overlap (**Fig 1B**). Differences in end vertebral span are due to differences in patient height. Initial data were epoched from -1 to +2 s relative to laser onset. Time-frequency decomposition was performed over a restricted window of -0.5 to 1 s to capture the primary evoked response while retaining sufficient pre-stimulus baseline for normalisation. All TFRs were normalised using a pre-stimulus baseline period (−0.5 to 0 s) with decibel conversion using the FieldTrip function ‘ft_freqbaseline’ (RRID: SCR_004849)[19], allowing for direct comparison of power changes across frequency bands. For visualisation, mild smoothing was applied across the frequency dimension using a 3-point weighted average (central weight 0.6, adjacent weights 0.2). Final plots were generated using ‘ft_singleplotTFR’ with custom parameters for each frequency range (5–30 Hz, 20–100 Hz, and 100–500 Hz).

**Figure 1.**
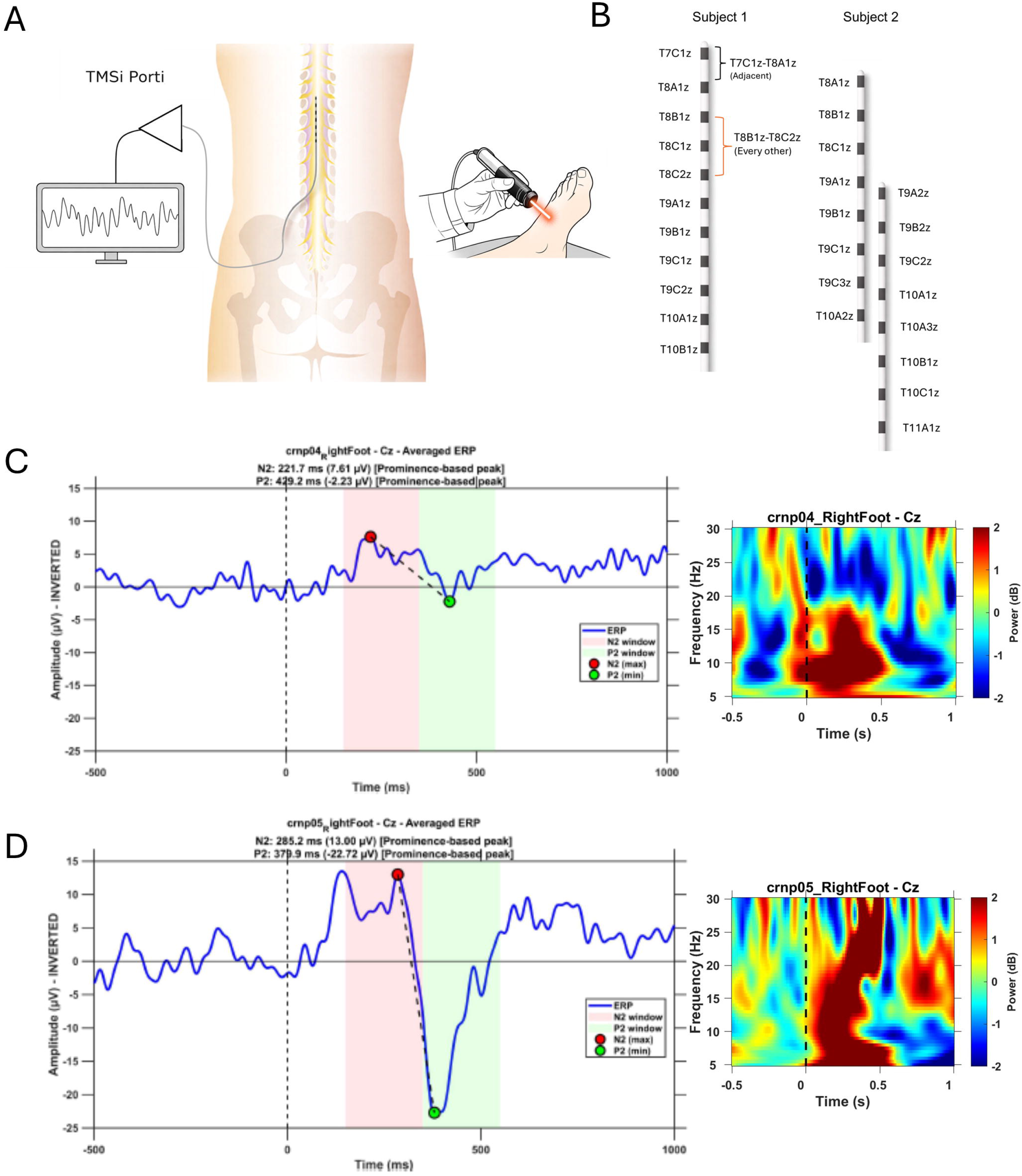
Cz event-related potentials and time–frequency representations during laser-evoked responses. **A)** Recording setup for externalised SFP acute pain experiment. **B)** Schematic of SCS electrode positions for each subject and channel names. For Subject 1, a single adjacent contact and every-other contact montage is shown as an example. Grand-average ERPs at Cz are shown for **(C)** Subject 1 (*crnp04*, top; n = 24 trials) and **(D)** Subject 2 (*crnp05*, bottom; n = 27 trials), time-locked to stimulus onset (0 ms; dashed line) on the left. Shaded regions indicate the predefined N2 (red) and P2 (green) LEP latency windows, with prominence-based peak latencies marked. Corresponding time–frequency representations (5–30 Hz) are shown on the right. ERPs demonstrate a N2–P2 complex, while time–frequency plots show low-frequency power modulation time-locked to the evoked response.

### Cluster-based permutation analysis of evoked-time frequency responses

eSFP responses to laser stimulation were analysed using non-parametric cluster-based permutation testing as implemented in the FieldTrip toolbox (MATLAB)[19]. All statistical analyses were performed separately for each subject, and no formal between-subject inference was attempted. Single-trial time–frequency representations were computed without baseline correction (FieldTrip ‘ft_freqanalysis’, cfg.keeptrials = ‘yes’) using the same Hanning-taper parameters as for visualisation (5 cycles per frequency, 4–35 Hz, 10 ms time steps, 20 s padding) over a time window of −0.5 to 1 s, with the frequency range expanded to 4–35 Hz to capture beta band (13–35 Hz) activity. For each trial, channel, and frequency, a baseline power estimate was obtained by averaging power across a pre-stimulus window. Post-stimulus power was then expressed as a decibel change relative to this trial-specific baseline (10 × log_10_[post-stimulus power/baseline power]), yielding a distribution of single-trial dB values at each time–frequency–channel point, where zero represents no change from baseline. A dependent-samples t-test against a zero-valued dataset of identical structure was used at the cluster level, corresponding to the null hypothesis that mean post-stimulus deviation from baseline equals zero. The permutation distribution was generated using 5,000 trial-wise sign flips, appropriate for within-subject data.

Clustering was performed jointly across time, frequency, and spatial dimensions. Clusters were formed by grouping adjacent samples exceeding an initial cluster-forming threshold of p < 0.05 or p < 0.01, and cluster-level significance was assessed using the maximum sum of suprathreshold t-values (maxsum) compared to the permutation distribution. Two-tailed testing was used throughout, with significant effects reported at p < 0.05 (corrected). Spatial adjacency was defined based on the rostro-caudal organisation of the epidural recordings. Bipolar channels were ordered along the long axis of each lead, with adjacency specified only between immediately neighbouring bipolar pairs along the same lead, forming a one-dimensional spatial chain. For Subject 2 (dual leads), each lead was treated as a separate chain with no inter-lead adjacency. This approach reflects the linear anatomical organisation of the spinal cord and favours detection of physiologically plausible, longitudinally contiguous responses, while limiting sensitivity to isolated contact-specific fluctuations. To assess robustness across analytic constraints, analyses were performed across multiple pre-stimulus baseline intervals (−500 to −100 ms; −400 to −100 ms; −300 to −100 ms), post-stimulus windows (early 0–200 ms vs. full 0–800 ms) to capture early stimulus-locked responses and temporally extended effects, frequency bands (low beta [13-20 Hz], and high beta [20-35 Hz]) and two bipolar montages (adjacent contact or every-other contact). Only spatially contiguous clusters (i.e. present in 2 or more contiguous bipolar channels) were considered significant. No statistical inference was performed across subjects; convergence of findings between subjects is reported qualitatively.

### Inter-trial coherence analysis

Inter-trial coherence (ITC) was computed to quantify the consistency of phase alignment across trials at each time–frequency point. Time–frequency decomposition was performed on single-trial bipolar signals using Hanning-tapered short-time Fourier transforms (FieldTrip ft_freqanalysis, method mtmconvol) with complex Fourier coefficients retained as output (cfg.output = ‘fourier’). Decomposition parameters matched those used for the power analysis: 4–35 Hz in 0.5 Hz steps, adaptive time windows of 5 cycles per frequency, and 10 ms time steps over a −0.5 to 1 s window. Individual trials were preserved (cfg.keeptrials = ‘yes’). For each channel, frequency, and time point, single-trial Fourier coefficients were normalised to unit magnitude to isolate phase information. The normalised complex values were summed across trials, and ITC was obtained as the absolute value of this sum divided by the number of trials, yielding values between 0 (no phase consistency across trials) and 1 (perfect phase alignment across trials). ITC was computed separately for each bipolar channel and subject and visualised as time–frequency maps aligned to stimulus onset. Given the limited trial numbers and the averaged nature of ITC, formal statistical testing was not performed. Instead, ITC patterns were examined descriptively alongside the power-based findings to assess whether observed oscillatory modulations were accompanied by stimulus-locked phase alignment.

## Results

Pain related oscillations generally report an early theta and gamma response (event-related synchronisation; ERS), followed by alpha and beta band event-related desynchronization (ERD) [21]. Thermal foot pain produced LEPs at Cz in both subjects as well as robust low-frequency power modulation (**Figure 1C-D**). **Figure 2** shows representative time-frequency plots for both subjects across three different frequency ranges (5-30 Hz, 20-100 Hz, and 100-500 Hz). As expected using cylindrical SCS leads with ring electrodes, at higher frequencies (**Fig 2C-F**) signals are dominated by a broadband increases consistent with non-neural (muscle) artefact, limiting further interpretation in these frequency ranges.

**Figure 2.**
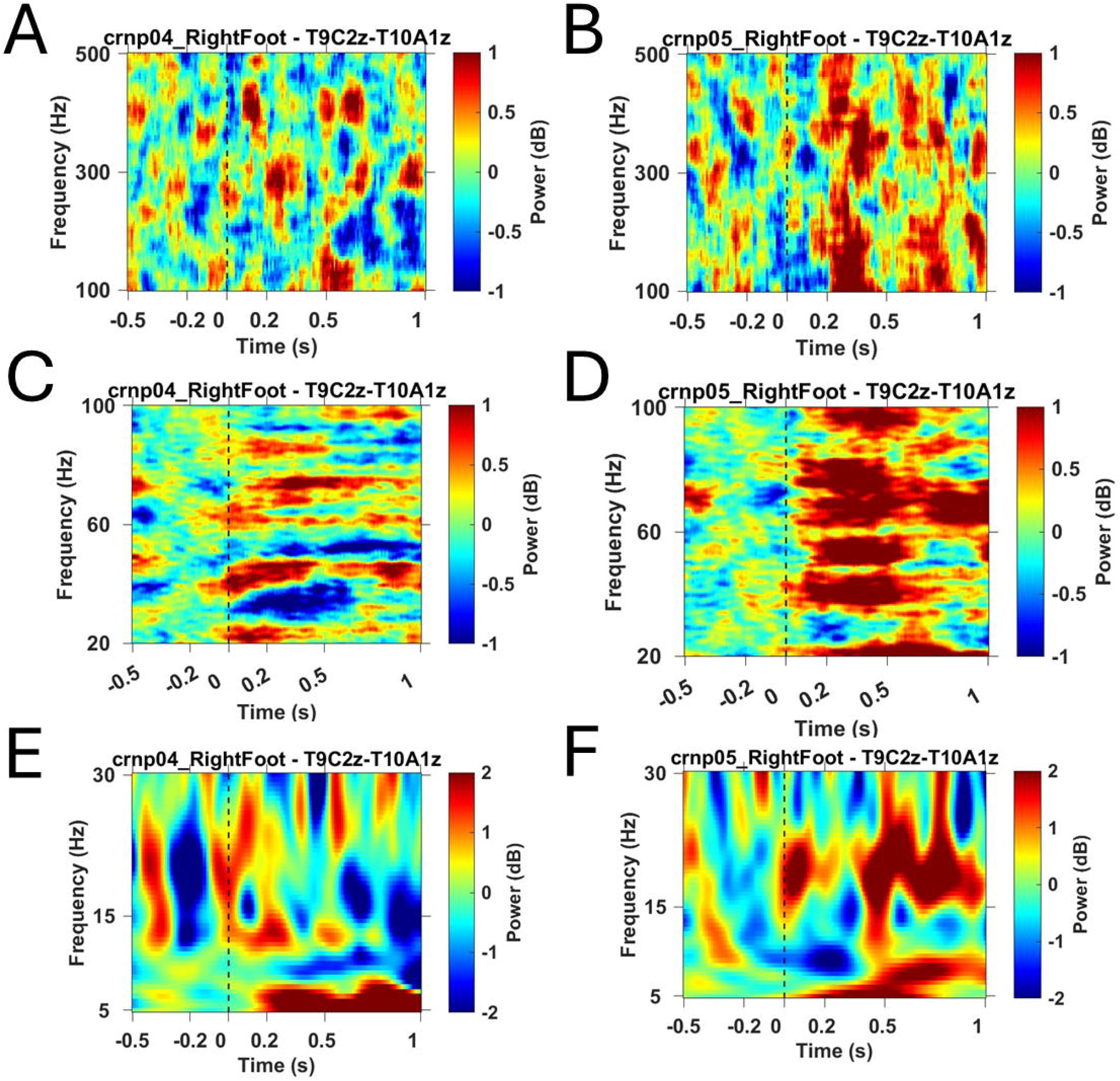
Time–frequency representations at rostrocaudally equivalent adjacent bipolar contacts in both subjects. Time–frequency plots from single bipolar channels from both Subject 1 (*crnp04*; **A**,**C**,**E**) and Subject 2 (*crnp05*; **B**,**D**,**F**) aligned to stimulus onset (0 s; dashed line). Power is shown as baseline-normalised change (dB). Top and middle rows depict high-frequency activity (100– 500 Hz and 20–100 Hz, respectively), which is dominated by broadband increases consistent with non-neural (muscle) artefact, limiting further interpretation in these frequency ranges. Bottom rows show low-frequency activity (5–30 Hz), which exhibits structured, time-locked modulation and forms the basis of subsequent analyses.

Figure 3 shows nociceptive stimulus evoked time-resolved changes in eSFP spectral power at 5-30 Hz (normalised to a -500 to 0 baseline) across adjacent contact bipolar channels in all subjects and leads. Cluster-based permutation analysis revealed consistent suppression of eSFP beta band activity following nociceptive stimulation across both subjects and all leads (**Supplementary Table 1**). In Subject 1 (crnp-04), using adjacent-contact bipolars, early (0-200ms) post-stimulus window showed significant low-beta suppression localised to more rostral contacts (T8A1z–T9C1z, cluster mass = −1881, *p* = 0.0006, spanning 0-200ms) and high beta suppression across most contacts (T7C1z–T10B1z, cluster mass = -7297, *p* = 0.0002, spanning 0-200ms). Using the 0–800 ms window showed similar early findings for low and high beta suppression, but with a more conservative cluster-forming threshold (p < 0.01) it also identified late clusters for high beta in middle contacts (T8C1z-T9B1z, cluster mass = -1682, *p* = 0.0014, spanning 570-760 ms). Every-other-contact bipolar referencing yielded similar results for early low- and high-beta suppression, but additionally detected late low-beta suppression at standard and more conservative clustering thresholds in rostral contacts (T8B1z-T9C1z, cluster mass = -988, *p* = 0.0046, spanning 620-800 ms).

**Figure 3.**
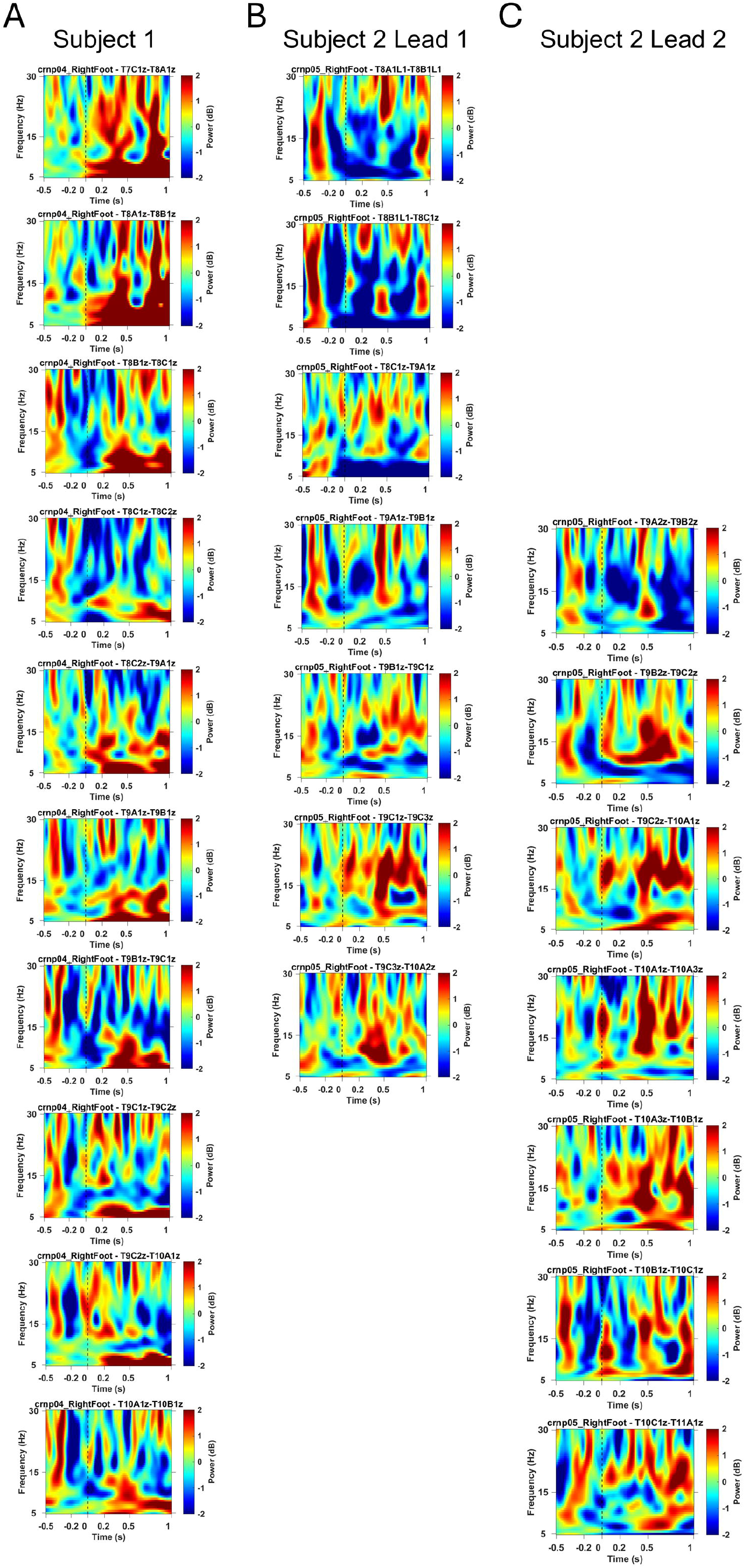
Rostrocaudally-resolved thermal pain-evoked time-frequency responses in adjacent contact bipolar epidural spinal field potentials. Time–frequency plots (5–30 Hz) are shown for overlapping adjacent contact bipolar channels arranged from rostral (top) to caudal (bottom) for **(A)** Subject 1 and **(B)** Subject 2 Lead 1 and **(C)** Subject 2 Lead 2, aligned to stimulus onset (t=0 s; vertical black dotted line). Power is expressed as baseline-normalised change (dB) and heat maps indicate high (red) and low (blue) signal power in each frequency band at different time points relative to painful stimulus. Across contacts, structured low-frequency modulation is observed with variability in magnitude and timing along the lead.

In Subject 2 (crnp-05), beta suppression again predominated but varied by lead. Adjacent contact analysis on the more rostral Lead 1, showed early low beta suppression involved middle channels (T8C1z–T9C1z; cluster mass = -537, *p* = 0.0136, spanning 0-200 ms). By contrast early high-beta suppression involved the entire lead for early (T8A1z-T10A2z; cluster mass = -5592, *p* = 0.0002, spanning 0-200ms) and almost the entire lead for late clusters (T8B1z-T10A2z; cluster mass = -1337, *p* = 0.0196, spanning 530-680ms), although only early clusters survive a more conservative cluster forming threshold (0.01). Every other contact analysis showed only early onset significant clusters for both low- and high-beta suppression. Adjacent contact analysis on the more caudal Lead 2 showed consistent low-beta suppression in the upper 2 channels only in both early (T9A2z-T9C2z, cluster mass = - 957, *p* = 0.001, spanning 0-200ms) and full windows (T9A2z-T9C2z, cluster mass = -1888, *p* = 0.0006, spanning 0-450ms) which did not survive a more conservative cluster threshold. By contrast, early high-beta suppression clusters formed across the entire electrode even at the most stringent cluster forming threshold of 0.01 (T9A2z-T11A1z; cluster mass = -3572, *p* = 0.0002, spanning 0-220ms). Every-other-contact analyses demonstrated a similar pattern.

Inspection of eSFP inter-trial coherence (ITC) for adjacent-contact bipolar channels revealed modest and spatially heterogeneous phase-locking across subjects and leads (**Figure 4**). Formal statistical testing of ITC was not performed at the single-subject level, as ITC is intrinsically defined across trials and available trial counts limited robust resampling-based inference; therefore, our observations are descriptive. In Subject 1 (**Fig 4A**), ITC increases were generally weak and spatially inconsistent across the rostral chain that showed the strongest low beta power suppression (T8A1z–T9A1z). Only subtle ITC elevations were observed across these adjacent bipolars, and these were most evident at later post-stimulus latencies rather than within the early 0–200 ms window where beta suppression was maximal.

**Figure 4.**
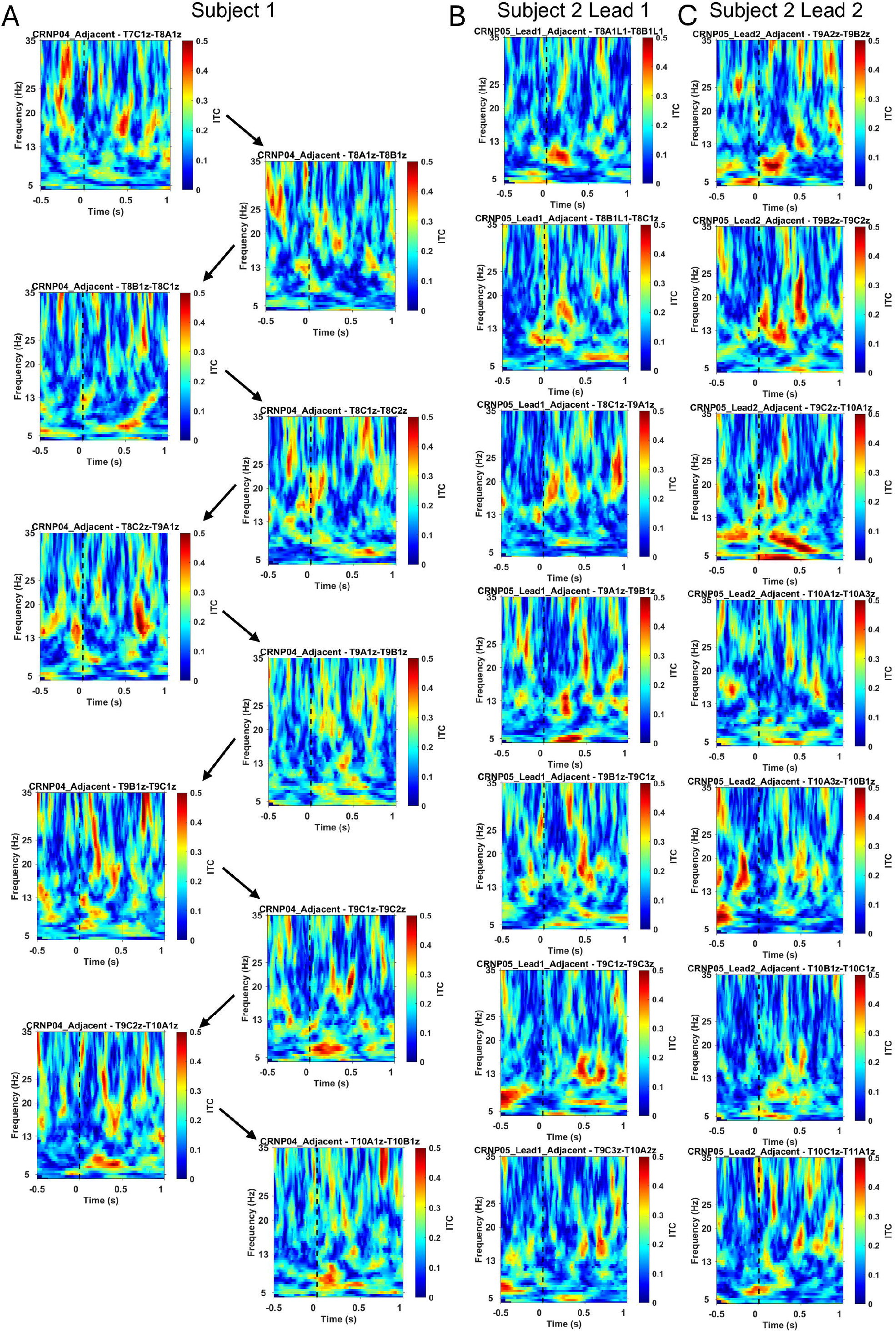
Rostrocaudally-resolved Inter-trial coherence (ITC) across adjacent bipolar spinal channels. Time–frequency representations of inter-trial coherence (ITC) for adjacent-contact bipolar recordings in **A)** Subject 1 and **(B)** Subject 2 Lead 1 and **(C)** Subject 2 Lead 2. Each panel shows ITC as a function of time relative to laser stimulus onset (vertical dashed line at 0 s) and frequency (5–35 Hz). Panels are arranged rostrocaudally from top to bottom across adjacent bipolar pairs. Warmer colours indicate higher phase consistency across trials. Across subjects, ITC increases were modest and spatially heterogeneous, with limited early phase alignment and more variable later increases, particularly in Subject 2. ITC values are shown without statistical thresholding, and panels are presented for descriptive comparison with power-based analyses.

In Subject 2, ITC structure was more pronounced but remained spatially and temporally dissociable from early power effects. On the more rostral Lead 1 (**Fig 4B**), ITC increases in T8A1L1–T9C1z were primarily observed at intermediate latencies (∼200–500 ms) and spanned beta frequencies. In contrast, the more caudal Lead 2 (**Fig 4C**), which demonstrated the strongest early high-beta suppression along most contacts, showed patchy early low-beta phase alignment within ∼0–200 ms across but more sustained later ITC increases in high beta extending into ∼300–700 ms. However, even in this lead, ITC increases were not uniformly aligned with the full spatial extent of early suppression and remained heterogeneous across adjacent channels.

In summary, early low-beta suppression within 0–200 ms was consistently observed along rostral bipolar chains across both subjects, while early high beta suppression was seen across the entire electrode. Adjacent-contact and every-other-contact configurations yielded similar clusters with small differences, indicating that early spatially distinct beta suppression represents the most consistent feature following nociceptive stimulus. Early beta suppression was not consistently accompanied by strong or spatially coherent ITC increases suggesting it likely reflects induced desynchronisation rather than a canonical phase-reset response.

## Discussion

Beta oscillations (13–35 Hz) are widely regarded as markers of sensorimotor network state, associated with tonic inhibition, maintenance of ongoing processing, and top–down control in cortical and subcortical systems [5,11]. Beta suppression is commonly linked to transient network reconfiguration and release from ongoing synchronisation [5,22]. In this study, we used eSFP recordings to characterise beta sub-band oscillatory responses to nociceptive stimulation in awake humans. By combining rostro-caudally-resolved bipolar recordings with cluster-based permutation analysis and explicit comparison of bipolar montage configurations, we identified reproducible beta sub-band suppression in two consecutively recorded subjects. Across analyses, beta-band suppression was the most robust feature, persisting across baseline definitions, temporal windows, and referencing schemes, and remaining detectable under conservative clustering thresholds (**Supplementary Table 1**). Applying stricter statistical criteria preferentially isolated early components of suppression, suggesting that rapid disruption of ongoing beta activity represents the most stable element of the spinal response to nociceptive stimuli. Notably, early beta suppression was robust at the power level but only weakly reflected in inter-trial coherence, indicating that nociceptive input primarily induces desynchronisation rather than a canonical phase-reset response. More structured ITC increases were observed at later latencies, suggesting that phase reorganisation may contribute to later stages of spinal network processing but is not required to explain the earliest beta dynamics. Notably, the rostral contacts showing the most consistent low-beta suppression corresponded to those providing the most effective analgesic coverage during the stimulation trial in both subjects (**Table 1**). Although a preliminary finding, this spatial concordance raises the possibility that low-beta suppression may index spinal networks therapeutically engaged in SCS.

Across both subjects, the most consistent effect was early beta suppression within 200 ms of nociceptive stimulation. While low-beta suppression dominated in rostral contacts, early high-beta suppression was consistently detectable across all leads. Beta suppression along contiguous adjacent bipolar chains may suggest that nociceptive response is organised longitudinally along segmental spinal networks rather than the dorsal root of the stimulated sensory dermatome. The co-occurrence of low- and high-beta suppression within early post-stimulus windows supports the view that nociceptive input produces rapid, multi-frequency disruption of ongoing oscillatory coordination. Although the functional significance of beta sub-bands remains debated, lower beta frequencies (13–20 Hz) are often linked to large-scale network state maintenance, whereas higher beta frequencies (20–35 Hz) have been associated with more local sensorimotor coupling [2,5,11]. The presence of both components in the early eSFP response may therefore reflect disruption of oscillatory coordination across multiple spatial scales.

Although laser stimulation provides a relatively selective nociceptive paradigm, caution is warranted in attributing early spinal beta suppression specifically to nociceptive processing as opposed to rapid spinal network modulation temporally associated with nociceptive input. Epidural recordings have previously demonstrated behavioural state–dependent spectral modulation [4,29], raising the possibility that the present effects reflect more general network reconfiguration. For example, the early 0–200 ms window could plausibly include spinal reflex-related activity triggered by Aδ afferent input. However, several features argue against a purely segmental or reflex interpretation. ITC was heterogeneous and did not show the strong phase alignment which might be expected from highly synchronised reflex responses [15,22,28] and the spatial predominance of rostral rather than caudal effects complicate a simple reflex interpretation. Notably, the observed suppression shares key features with cortical beta event-related desynchronisation, including rapid onset and dominance at the power level over phase locking [21,22]. This raises the possibility that nociceptive input induces a conserved desynchronisation response across the nervous system, reflecting transient disruption of ongoing network synchrony rather than a strictly nociception-specific or reflex-bound phenomenon [18,23]. At the spinal level, this may reflect disruption of ongoing oscillatory coordination within dorsal horn and propriospinal circuits following nociceptive input [12] though it remains unclear why temporally synchronous rostro-caudal differences in beta sub-band responses would arise.

The magnitude and spatial coherence of beta suppression were affected by baseline selection and post-stimulus analysis window more than the exact bipolar montage. Differences in non-overlapping adjacent-contact and overlapping every-other-contact configurations likely reflect the mixing of signals across non-contiguous segments. Similarly, increasing clustering stringency (from cluster forming threshold *p* < 0.05 to *p* < 0.01) preferentially eliminated later and spatially extended effects while preserving early contiguous suppression, reinforcing the robustness of early beta dynamics. These findings highlight key considerations for analysing eSFPs as spatial referencing strongly influences the apparent topology of oscillatory effects, and conservative statistical thresholds preferentially isolate temporally focal, spatially contiguous responses.

Several limitations warrant consideration beyond the small number of subjects in this preliminary study. The number of trials was constrained by the clinical recording context, pain ratings were not recorded on a trial-by-trial basis and non-painful control stimuli were not delivered, precluding direct assessment of the relationship between spinal oscillatory dynamics and subjective pain perception. We did not record paraspinal electromyography preventing definitive exclusion of muscle contributions, although the spectral specificity, temporal structure, and montage dependence of the observed beta effects argue against a predominantly myogenic origin which is seen as high frequency power increase [6,17]. Both subjects were also receiving anti-neuropathic medications, which are known to modulate nociceptive processing and may therefore influence oscillatory dynamics [10]. The use of adaptive time–frequency windows for TFRs prioritised spectral precision over temporal resolution and may have reduced sensitivity to very brief early modulation. Finally, electrode geometry and electrode–cord coupling may influence spatial sampling, and future recordings obtained with alternative electrode designs (e.g. paddles) may provide improved resolution of local spinal generators.

Our preliminary findings provide evidence that nociceptive stimulation can induce rapid and spatially distinct suppression of spinal beta sub-band activity in humans. The persistence of early suppression across subjects, leads, and analyses suggests that spinal beta dynamics are consistent with a physiological response to nociceptive stimulation. From a translational perspective, the ability to detect early evoked oscillatory signatures using clinically implanted epidural electrodes raises the possibility that spinal beta activity could serve as a biomarker of spinal network state or detect therapeutically relevant spinal networks to inform adaptive neuromodulation strategies.

## Data Availability

All data produced in the present study are available upon reasonable request to the authors

## Acknowledgements

R.S.S was personally supported by an NIHR CL award (CL-2021-16-1502) and an RCS England Pump Priming grant funded by Saven Research & Development Fund. A.M.N was supported by a Saluda Medical Research Grant. We thank Stoke Mandeville Spinal Research Charity for funding scientific equipment.

## Data availability

Data is available on reasonable request.

## Declaration of generative AI and AI-assisted technologies in the writing process

During the preparation of the draft, the first author (RS) used ChatGPT and Gemini to improve readability and language fluency during the writing process. After using the AI-assisted technologies, the author reviewed and edited the content and took full responsibility for the manuscript.

