## Supplemental data for "Nociceptive stimulation suppresses spinal beta oscillations in awake human epidural recordings"

Supplementary Methods

*Laser-thermal stimuli recordings*

On their end of trial visit, the trial SCS electrodes were disconnected from their external stimulator and connected to an electrophysiology recording system using custom-made cables for passive recording as described previously [29]. Both recordings were performed on postoperative day 13 when epidural air and blood have generally reabsorbed. The experimental schematic is shown in Figure 1A. The subjects were lying down for the duration of the experiment to limit postural muscle artefact and maximise epidural electrode proximity to the spinal cord. A TMSi Porti amplifier (2048 Hz sampling rate, left forearm ground electrode; TMS International, Netherlands) was used to simultaneously record monopolar spinal field potentials (SFPs), EEG (Cz, nose, T3,T4), and the timing of triggers for thermal laser stimulation using custom written software developed using the C programming language (Figure 1A). Aδ-LEP components were recorded through disk electrodes from the scalp (Cz referenced to the nose). Nociceptive-specific radiant-heat stimuli were generated by an infrared neodymium yttrium aluminum perovskite (Nd:YAP) laser with fibre-optic guidance (1340 nm wavelength, 5 mm beam diameter, 5 ms pulse duration; Stimul 1340, DEKA, Italy). At this wavelength, laser pulses directly activate nociceptive terminals in the most superficial skin layers [3,9]. Laser pulses were directed on an area ($5 \times 5$ $cm^2$) on the dorsal aspect of the foot. A red beam pointed to the target area to be stimulated. After each stimulus, the target of the laser beam was shifted by at least 1 cm in a random direction to allow for passive skin cooling and avoid nociceptor fatigue or sensitization. To keep attention stable subjects were asked to mentally count the number of stimuli. Power of laser stimuli were increased in 0.25J or 0.5J increments (from a starting power of 3.0J) until pain scores reported were at least 4 (clearly painful) at which point recordings started. Interstimulus interval was varied on purpose to avoid predictability but was not less than 10 seconds. Individual pain score per stimulus was not recorded. At the end of the recording their trial SCS cables were removed in an aseptic fashion and the exit site dressed.

*Laser Evoked Potential Analysis*

LEPs were extracted from the Cz EEG channel using a time window of −1 to 2 seconds relative to laser stimulus onset ($t = 0$). In Subject 1, Cz was re-referenced to the average of T3 + T4, whereas in Subject 2 Cz was re-referenced to the Nose electrode. The signal underwent bandpass filtering between 0.5 to 30 Hz. Trials were averaged across epochs to generate individual event-related potentials (ERPs) for each subject. Based on their temporal characteristics, we identified the key LEP components on EEG: N2 (maximum negative peak in the 150–350 ms post-stimulus window) and P2 (maximum positive peak in the 350–550 ms post-stimulus window). Because the standard scalp polarity convention represents N2 as a negative deflection and P2 as a positive deflection, the Cz signal was inverted before the automated peak detection so that the N2 appeared as a local maximum and the P2 as a local minimum. LEP components were extracted from each subject's trial-averaged ERP using an automated prominence-based peak detection method (MATLAB findpeaks). Within the N2 search window (150–350 ms post-stimulus), all local positive peaks (maxima) in the inverted signal were identified and stored as potential candidates, provided they exceeded a minimum prominence of 0.5 times the standard deviation of the signal within that window and a minimum peak width of 3 samples. When multiple candidate peaks met these criteria, each candidate's amplitude and prominence were independently normalised to z-scores across the set, and a score was computed as the sum of the normalised amplitude and 0.5 times the normalised prominence, such that amplitude contributed more than prominence to the final selection. The candidate with the highest score was selected as the N2 peak. The P2 component was identified as the most prominent negative peak (minimum) in the inverted signal within 350–550 ms using the same prominence and width thresholds. If no peaks met these thresholds, the absolute maximum or minimum within the respective window was used as a fallback; however, the prominence-based method successfully extracted both N2 and P2 components in both subjects without requiring fallback detection as shown in Figure 1. Peak latency and amplitude were extracted for each component.

| Subject | Bipolar montage | Baseline | Post-stim | Beta band | P Value | Cluster Stat | Cluster freq (Hz) | Time (ms) | Cluster-α | Channels |
|---|---|---|---|---|---|---|---|---|---|---|
| CRNP-04 | Adjacent | -300 to -100 ms | 0 to 800 ms | 13-20Hz | 0.0394 | -827.98 | 13.0–20.0 | 0–170 | 0.05 | T8A1z-T8B1z, T8B1z-T8C1z, T8C1z-T8C2z |
| CRNP-04 | Adjacent | -400 to -100 ms | 0 to 800 ms | 13-20Hz | 0.0008 | -2707.73 | 13.0–20.0 | 0–420 | 0.05 | T8A1z-T8B1z, T8B1z-T8C1z, T8C1z-T8C2z, T8C2z-T9A1z, T9A1z-T9B1z, T9B1z-T9C1z |
| CRNP-04 | Adjacent | -400 to -100 ms | 0 to 800 ms | 13-20Hz | 0.0026 | -2281.23 | 13.0–20.0 | 400–800 | 0.05 | T7C1z-T8A1z, T8A1z-T8B1z, T8B1z-T8C1z, T8C1z-T8C2z, T8C2z-T9A1z, T9A1z-T9B1z, T9B1z-T9C1z |
| CRNP-04 | Adjacent | -500 to -100 ms | 0 to 800 ms | 13-20Hz | 0.001 | -3279.03 | 13.0–20.0 | 0–530 | 0.05 | T8A1z-T8B1z, T8B1z-T8C1z, T8C1z-T8C2z, T8C2z-T9A1z, T9A1z-T9B1z, T9B1z-T9C1z, T9C1z-T9C2z |
| CRNP-04 | Adjacent | -500 to -100 ms | 0 to 800 ms | 13-20Hz | 0.0018 | -2682.43 | 13.0–20.0 | 400–800 | 0.05 | T7C1z-T8A1z, T8A1z-T8B1z, T8B1z-T8C1z, T8C1z-T8C2z, T8C2z-T9A1z, T9A1z-T9B1z, T9B1z-T9C1z, T9C1z-T9C2z |
| CRNP-04 | Adjacent | -300 to -100 ms | 0 to 200 ms | 13-20Hz | 0.0034 | -827.98 | 13.0–20.0 | 0–170 | 0.05 | T8A1z-T8B1z, T8B1z-T8C1z, T8C1z-T8C2z |
| CRNP-04 | Adjacent | -400 to -100 ms | 0 to 200 ms | 13-20Hz | 0.0006 | -1880.62 | 13.0–20.0 | 0–200 | 0.05 | T8A1z-T8B1z, T8B1z-T8C1z, T8C1z-T8C2z, T8C2z-T9A1z, T9A1z-T9B1z, T9B1z-T9C1z |
| CRNP-04 | Adjacent | -500 to -100 ms | 0 to 200 ms | 13-20Hz | 0.0004 | -2234.05 | 13.0–20.0 | 0–200 | 0.05 | T8A1z-T8B1z, T8B1z-T8C1z, T8C1z-T8C2z, T8C2z-T9A1z, T9A1z-T9B1z, T9B1z-T9C1z |
| CRNP-04 | Adjacent | -500 to -100 ms | 0 to 800 ms | 13-20Hz | 0.0224 | -434.32 | 13.0–20.0 | 0–160 | 0.01 | T8B1z-T8C1z, T8C1z-T8C2z, T8C2z-T9A1z |
| CRNP-04 | Adjacent | -400 to -100 ms | 0 to 200 ms | 13-20Hz | 0.0218 | -230.87 | 15.0–20.0 | 30–110 | 0.01 | T8A1z-T8B1z, T8B1z-T8C1z |
| CRNP-04 | Adjacent | -500 to -100 ms | 0 to 200 ms | 13-20Hz | 0.0044 | -434.32 | 13.0–20.0 | 0–160 | 0.01 | T8B1z-T8C1z, T8C1z-T8C2z, T8C2z-T9A1z |
| CRNP-04 | Adjacent | -500 to -100 ms | 0 to 200 ms | 13-20Hz | 0.0248 | -236.36 | 15.0–20.0 | 40–110 | 0.01 | T8A1z-T8B1z, T8B1z-T8C1z |
| CRNP-04 | Adjacent | -300 to -100 ms | 0 to 800 ms | 20-35Hz | 0.0002 | -12517.93 | 20.0–35.0 | 0–580 | 0.05 | T7C1z-T8A1z, T8A1z-T8B1z, T8B1z-T8C1z, T8C1z-T8C2z, T8C2z-T9A1z, T9A1z-T9B1z, T9B1z-T9C1z, T9C1z-T9C2z, T9C2z-T10A1z, T10A1z-T10B1z |
| CRNP-04 | Adjacent | -300 to -100 ms | 0 to 800 ms | 20-35Hz | 0.017 | -1772.93 | 20.0–35.0 | 550–800 | 0.05 | T8A1z-T8B1z, T8B1z-T8C1z, T8C1z-T8C2z, T8C2z-T9A1z |
| CRNP-04 | Adjacent | -400 to -100 ms | 0 to 800 ms | 20-35Hz | 0.0002 | -23466.86 | 20.0–35.0 | 0–800 | 0.05 | T7C1z-T8A1z, T8A1z-T8B1z, T8B1z-T8C1z, T8C1z-T8C2z, T8C2z-T9A1z, T9A1z-T9B1z, T9B1z-T9C1z, T9C1z-T9C2z, T9C2z-T10A1z, T10A1z-T10B1z |
| CRNP-04 | Adjacent | -500 to -100 ms | 0 to 800 ms | 20-35Hz | 0.0002 | -28475.86 | 20.0–35.0 | 0–800 | 0.05 | T7C1z-T8A1z, T8A1z-T8B1z, T8B1z-T8C1z, T8C1z-T8C2z, T8C2z-T9A1z, T9A1z-T9B1z, T9B1z-T9C1z, T9C1z-T9C2z, T9C2z-T10A1z, T10A1z-T10B1z |
| CRNP-04 | Adjacent | -300 to -100 ms | 0 to 200 ms | 20-35Hz | 0.0002 | -4438.25 | 20.0–35.0 | 0–200 | 0.05 | T7C1z-T8A1z, T8A1z-T8B1z, T8B1z-T8C1z, T8C1z-T8C2z, T8C2z-T9A1z, T9A1z-T9B1z, T9B1z-T9C1z, T9C1z-T9C2z, T9C2z-T10A1z, T10A1z-T10B1z |
| CRNP-04 | Adjacent | -400 to -100 ms | 0 to 200 ms | 20-35Hz | 0.0002 | -7296.92 | 20.0–35.0 | 0–200 | 0.05 | T7C1z-T8A1z, T8A1z-T8B1z, T8B1z-T8C1z, T8C1z-T8C2z, T8C2z-T9A1z, T9A1z-T9B1z, T9B1z-T9C1z, T9C1z-T9C2z, T9C2z-T10A1z, T10A1z-T10B1z |
| CRNP-04 | Adjacent | -500 to -100 ms | 0 to 200 ms | 20-35Hz | 0.0002 | -8964.2 | 20.0–35.0 | 0–200 | 0.05 | T7C1z-T8A1z, T8A1z-T8B1z, T8B1z-T8C1z, T8C1z-T8C2z, T8C2z-T9A1z, T9A1z-T9B1z, T9B1z-T9C1z, T9C1z-T9C2z, T9C2z-T10A1z, T10A1z-T10B1z |
| CRNP-04 | Adjacent | -300 to -100 ms | 0 to 800 ms | 20-35Hz | 0.0002 | -1765.45 | 20.0–35.0 | 0–150 | 0.01 | T8A1z-T8B1z, T8B1z-T8C1z, T8C1z-T8C2z, T8C2z-T9A1z |
| CRNP-04 | Adjacent | -300 to -100 ms | 0 to 800 ms | 20-35Hz | 0.0012 | -1186.57 | 26.5–35.0 | 370–510 | 0.01 | T9B1z-T9C1z, T9C1z-T9C2z, T9C2z-T10A1z, T10A1z-T10B1z |
| CRNP-04 | Adjacent | -300 to -100 ms | 0 to 800 ms | 20-35Hz | 0.0064 | -767.91 | 20.0–35.0 | 200–330 | 0.01 | T8A1z-T8B1z, T8B1z-T8C1z, T8C1z-T8C2z |
| CRNP-04 | Adjacent | -300 to -100 ms | 0 to 800 ms | 20-35Hz | 0.0094 | -682.54 | 20.0–27.0 | 570–750 | 0.01 | T8C1z-T8C2z, T8C2z-T9A1z |
| CRNP-04 | Adjacent | -300 to -100 ms | 0 to 800 ms | 20-35Hz | 0.0108 | -662.13 | 29.0–35.0 | 210–310 | 0.01 | T9C1z-T9C2z, T9C2z-T10A1z, T10A1z-T10B1z |
| CRNP-04 | Adjacent | -400 to -100 ms | 0 to 800 ms | 20-35Hz | 0.0002 | -3107.68 | 20.0–35.0 | 140–430 | 0.01 | T8A1z-T8B1z, T8B1z-T8C1z, T8C1z-T8C2z, T8C2z-T9A1z |
| CRNP-04 | Adjacent | -400 to -100 ms | 0 to 800 ms | 20-35Hz | 0.0004 | -2519.89 | 20.0–35.0 | 0–160 | 0.01 | T8A1z-T8B1z, T8B1z-T8C1z, T8C1z-T8C2z, T8C2z-T9A1z |
| CRNP-04 | Adjacent | -400 to -100 ms | 0 to 800 ms | 20-35Hz | 0.0004 | -2108.69 | 20.0–35.0 | 360–510 | 0.01 | T9A1z-T9B1z, T9B1z-T9C1z, T9C1z-T9C2z, T9C2z-T10A1z, T10A1z-T10B1z |
| CRNP-04 | Adjacent | -400 to -100 ms | 0 to 800 ms | 20-35Hz | 0.001 | -1406.65 | 20.0–35.0 | 560–760 | 0.01 | T8C1z-T8C2z, T8C2z-T9A1z, T9A1z-T9B1z |
| CRNP-04 | Adjacent | -400 to -100 ms | 0 to 800 ms | 20-35Hz | 0.0088 | -780.29 | 28.5–35.0 | 210–310 | 0.01 | T9C1z-T9C2z, T9C2z-T10A1z, T10A1z-T10B1z |
| CRNP-04 | Adjacent | -500 to -100 ms | 0 to 800 ms | 20-35Hz | 0.0002 | -8134.42 | 20.0–35.0 | 0–430 | 0.01 | T7C1z-T8A1z, T8A1z-T8B1z, T8B1z-T8C1z, T8C1z-T8C2z, T8C2z-T9A1z, T9A1z-T9B1z, T9B1z-T9C1z, T9C1z-T9C2z, T9C2z-T10A1z, T10A1z-T10B1z |
| CRNP-04 | Adjacent | -500 to -100 ms | 0 to 800 ms | 20-35Hz | 0.0002 | -2376.5 | 20.0–35.0 | 360–520 | 0.01 | T9A1z-T9B1z, T9B1z-T9C1z, T9C1z-T9C2z, T9C2z-T10A1z, T10A1z-T10B1z |
| CRNP-04 | Adjacent | -500 to -100 ms | 0 to 800 ms | 20-35Hz | 0.0014 | -1681.91 | 20.0–35.0 | 570–760 | 0.01 | T8C1z-T8C2z, T8C2z-T9A1z, T9A1z-T9B1z |
| CRNP-04 | Adjacent | -500 to -100 ms | 0 to 800 ms | 20-35Hz | 0.0062 | -930.98 | 27.5–35.0 | 200–310 | 0.01 | T9C1z-T9C2z, T9C2z-T10A1z, T10A1z-T10B1z |
| CRNP-04 | Adjacent | -300 to -100 ms | 0 to 200 ms | 20-35Hz | 0.0002 | -1765.45 | 20.0–35.0 | 0–150 | 0.01 | T8A1z-T8B1z, T8B1z-T8C1z, T8C1z-T8C2z, T8C2z-T9A1z |
| CRNP-04 | Adjacent | -400 to -100 ms | 0 to 200 ms | 20-35Hz | 0.0002 | -2519.89 | 20.0–35.0 | 0–160 | 0.01 | T8A1z-T8B1z, T8B1z-T8C1z, T8C1z-T8C2z, T8C2z-T9A1z |
| CRNP-04 | Adjacent | -400 to -100 ms | 0 to 200 ms | 20-35Hz | 0.0154 | -392.7 | 22.5–35.0 | 0–50 | 0.01 | T9C1z-T9C2z, T9C2z-T10A1z, T10A1z-T10B1z |
| CRNP-04 | Adjacent | -500 to -100 ms | 0 to 200 ms | 20-35Hz | 0.0002 | -4444.8 | 20.0–35.0 | 0–200 | 0.01 | T7C1z-T8A1z, T8A1z-T8B1z, T8B1z-T8C1z, T8C1z-T8C2z, T8C2z-T9A1z, T9A1z-T9B1z, T9B1z-T9C1z, T9C1z-T9C2z, T9C2z-T10A1z, T10A1z-T10B1z |
| CRNP-04 | Every Other | -300 to -100 ms | 0 to 800 ms | 13-20Hz | 0.0214 | -1286.7 | 13.0–20.0 | 560–800 | 0.05 | T7C1z-T8B1z, T8A1z-T8C1z, T8B1z-T8C2z, T8C1z-T9A1z, T8C2z-T9B1z, T9A1z-T9C1z |
| CRNP-04 | Every Other | -400 to -100 ms | 0 to 800 ms | 13-20Hz | 0.002 | -3079.17 | 13.0–20.0 | 490–800 | 0.05 | T7C1z-T8B1z, T8A1z-T8C1z, T8B1z-T8C2z, T8C1z-T9A1z, T8C2z-T9B1z, T9A1z-T9C1z |
| CRNP-04 | Every Other | -400 to -100 ms | 0 to 800 ms | 13-20Hz | 0.0228 | -1430.21 | 13.0–20.0 | 0–180 | 0.05 | T8A1z-T8C1z, T8B1z-T8C2z, T8C1z-T9A1z, T8C2z-T9B1z, T9A1z-T9C1z, T9B1z-T9C2z |
| CRNP-04 | Every Other | -500 to -100 ms | 0 to 800 ms | 13-20Hz | 0.0002 | -6619.07 | 13.0–20.0 | 270–800 | 0.05 | T7C1z-T8B1z, T8A1z-T8C1z, T8B1z-T8C2z, T8C1z-T9A1z, T8C2z-T9B1z, T9A1z-T9C1z, T9B1z-T9C2z, T9C1z-T10A1z, T9C2z-T10B1z |
| CRNP-04 | Every Other | -500 to -100 ms | 0 to 800 ms | 13-20Hz | 0.0098 | -2214.36 | 13.0–20.0 | 0–230 | 0.05 | T7C1z-T8B1z, T8A1z-T8C1z, T8B1z-T8C2z, T8C1z-T9A1z, T8C2z-T9B1z, T9A1z-T9C1z, T9B1z-T9C2z |
| CRNP-04 | Every Other | -300 to -100 ms | 0 to 200 ms | 13-20Hz | 0.0242 | -598.59 | 13.0–20.0 | 0–150 | 0.05 | T8B1z-T8C2z, T8C1z-T9A1z, T8C2z-T9B1z |
| CRNP-04 | Every Other | -400 to -100 ms | 0 to 200 ms | 13-20Hz | 0.0026 | -1430.21 | 13.0–20.0 | 0–180 | 0.05 | T8A1z-T8C1z, T8B1z-T8C2z, T8C1z-T9A1z, T8C2z-T9B1z, T9A1z-T9C1z, T9B1z-T9C2z |
| CRNP-04 | Every Other | -400 to -100 ms | 0 to 200 ms | 13-20Hz | 0.027 | -636.35 | 14.0–20.0 | 0–200 | 0.05 | T9C1z-T10A1z, T9C2z-T10B1z |
| CRNP-04 | Every Other | -500 to -100 ms | 0 to 200 ms | 13-20Hz | 0.0012 | -2188.19 | 13.0–20.0 | 0–200 | 0.05 | T7C1z-T8B1z, T8A1z-T8C1z, T8B1z-T8C2z, T8C1z-T9A1z, T8C2z-T9B1z, T9A1z-T9C1z, T9B1z-T9C2z |
| CRNP-04 | Every Other | -500 to -100 ms | 0 to 200 ms | 13-20Hz | 0.0154 | -887.41 | 13.5–20.0 | 0–200 | 0.05 | T9C1z-T10A1z, T9C2z-T10B1z |
| CRNP-04 | Every Other | -400 to -100 ms | 0 to 800 ms | 13-20Hz | 0.0132 | -664.52 | 13.0–20.0 | 660–800 | 0.01 | T8B1z-T8C2z, T8C1z-T9A1z, T8C2z-T9B1z, T9A1z-T9C1z |
| CRNP-04 | Every Other | -400 to -100 ms | 0 to 800 ms | 13-20Hz | 0.0134 | -636.58 | 13.0–20.0 | 0–150 | 0.01 | T8B1z-T8C2z, T8C1z-T9A1z, T8C2z-T9B1z |
| CRNP-04 | Every Other | -400 to -100 ms | 0 to 800 ms | 13-20Hz | 0.0418 | -386.38 | 16.0–20.0 | 600–720 | 0.01 | T7C1z-T8B1z, T8A1z-T8C1z, T8B1z-T8C2z, T8C1z-T9A1z, T8C2z-T9B1z |
| CRNP-04 | Every Other | -500 to -100 ms | 0 to 800 ms | 13-20Hz | 0.0046 | -987.92 | 13.0–20.0 | 620–800 | 0.01 | T8B1z-T8C2z, T8C1z-T9A1z, T8C2z-T9B1z, T9A1z-T9C1z |
| CRNP-04 | Every Other | -500 to -100 ms | 0 to 800 ms | 13-20Hz | 0.0058 | -887.89 | 13.0–20.0 | 0–150 | 0.01 | T8B1z-T8C2z, T8C1z-T9A1z, T8C2z-T9B1z |
| CRNP-04 | Every Other | -500 to -100 ms | 0 to 800 ms | 13-20Hz | 0.0122 | -672.61 | 15.5–20.0 | 600–750 | 0.01 | T7C1z-T8B1z, T8A1z-T8C1z, T8B1z-T8C2z, T8C1z-T9A1z, T8C2z-T9B1z |
| CRNP-04 | Every Other | -500 to -100 ms | 0 to 800 ms | 13-20Hz | 0.0412 | -383.22 | 14.5–20.0 | 0–150 | 0.01 | T9C1z-T10A1z, T9C2z-T10B1z |
| CRNP-04 | Every Other | -400 to -100 ms | 0 to 200 ms | 13-20Hz | 0.002 | -636.58 | 13.0–20.0 | 0–150 | 0.01 | T8B1z-T8C2z, T8C1z-T9A1z, T8C2z-T9B1z |
| CRNP-04 | Every Other | -500 to -100 ms | 0 to 200 ms | 13-20Hz | 0.0014 | -887.89 | 13.0–20.0 | 0–150 | 0.01 | T8B1z-T8C2z, T8C1z-T9A1z, T8C2z-T9B1z |

| Subject | Bipolar montage | Baseline | Post-stim | Beta band | P Value | Cluster Stat | Cluster freq (Hz) | Time (ms) | Cluster-α | Channels |
|---|---|---|---|---|---|---|---|---|---|---|
| CRNP-04 | Every Other | -500 to -100 ms | 0 to 200 ms | 13-20Hz | 0.0096 | -383.22 | 14.5–20.0 | 0–150 | 0.01 | T9C1z-T10A1z, T9C2z-T10B1z |
| CRNP-04 | Every Other | -300 to -100 ms | 0 to 800 ms | 20-35Hz | 0.0002 | -12292.27 | 20.0–35.0 | 0–610 | 0.05 | T7C1z-T8B1z, T8A1z-T8C1z, T8B1z-T8C2z, T8C1z-T9A1z, T8C2z-T9B1z, T9A1z-T9C1z, T9B1z-T9C2z, T9C1z-T10A1z, T9C2z-T10B1z |
| CRNP-04 | Every Other | -400 to -100 ms | 0 to 800 ms | 20-35Hz | 0.0002 | -21481.35 | 20.0–35.0 | 0–800 | 0.05 | T7C1z-T8B1z, T8A1z-T8C1z, T8B1z-T8C2z, T8C1z-T9A1z, T8C2z-T9B1z, T9A1z-T9C1z, T9B1z-T9C2z, T9C1z-T10A1z, T9C2z-T10B1z |
| CRNP-04 | Every Other | -500 to -100 ms | 0 to 800 ms | 20-35Hz | 0.0002 | -27577.26 | 20.0–35.0 | 0–800 | 0.05 | T7C1z-T8B1z, T8A1z-T8C1z, T8B1z-T8C2z, T8C1z-T9A1z, T8C2z-T9B1z, T9A1z-T9C1z, T9B1z-T9C2z, T9C1z-T10A1z, T9C2z-T10B1z |
| CRNP-04 | Every Other | -300 to -100 ms | 0 to 200 ms | 20-35Hz | 0.0002 | -4858.93 | 20.0–35.0 | 0–200 | 0.05 | T7C1z-T8B1z, T8A1z-T8C1z, T8B1z-T8C2z, T8C1z-T9A1z, T8C2z-T9B1z, T9A1z-T9C1z, T9B1z-T9C2z, T9C1z-T10A1z, T9C2z-T10B1z |
| CRNP-04 | Every Other | -400 to -100 ms | 0 to 200 ms | 20-35Hz | 0.0002 | -7166.32 | 20.0–35.0 | 0–200 | 0.05 | T7C1z-T8B1z, T8A1z-T8C1z, T8B1z-T8C2z, T8C1z-T9A1z, T8C2z-T9B1z, T9A1z-T9C1z, T9B1z-T9C2z, T9C1z-T10A1z, T9C2z-T10B1z |
| CRNP-04 | Every Other | -500 to -100 ms | 0 to 200 ms | 20-35Hz | 0.0002 | -8875.43 | 20.0–35.0 | 0–200 | 0.05 | T7C1z-T8B1z, T8A1z-T8C1z, T8B1z-T8C2z, T8C1z-T9A1z, T8C2z-T9B1z, T9A1z-T9C1z, T9B1z-T9C2z, T9C1z-T10A1z, T9C2z-T10B1z |
| CRNP-04 | Every Other | -300 to -100 ms | 0 to 800 ms | 20-35Hz | 0.0002 | -1614.95 | 23.5–35.0 | 0–110 | 0.01 | T8A1z-T8C1z, T8B1z-T8C2z, T8C1z-T9A1z, T8C2z-T9B1z |
| CRNP-04 | Every Other | -300 to -100 ms | 0 to 800 ms | 20-35Hz | 0.0016 | -953.08 | 24.5–35.0 | 200–410 | 0.01 | T8A1z-T8C1z, T8B1z-T8C2z, T8C1z-T9A1z, T8C2z-T9B1z |
| CRNP-04 | Every Other | -300 to -100 ms | 0 to 800 ms | 20-35Hz | 0.0118 | -611.53 | 24.5–35.0 | 430–510 | 0.01 | T8C2z-T9B1z, T9A1z-T9C1z, T9B1z-T9C2z, T9C1z-T10A1z, T9C2z-T10B1z |
| CRNP-04 | Every Other | -400 to -100 ms | 0 to 800 ms | 20-35Hz | 0.0002 | -3197.32 | 20.0–35.0 | 0–180 | 0.01 | T7C1z-T8B1z, T8A1z-T8C1z, T8B1z-T8C2z, T8C1z-T9A1z, T8C2z-T9B1z, T9A1z-T9C1z, T9B1z-T9C2z |
| CRNP-04 | Every Other | -400 to -100 ms | 0 to 800 ms | 20-35Hz | 0.0002 | -2117.31 | 23.0–35.0 | 200–410 | 0.01 | T7C1z-T8B1z, T8A1z-T8C1z, T8B1z-T8C2z, T8C1z-T9A1z, T8C2z-T9B1z |
| CRNP-04 | Every Other | -400 to -100 ms | 0 to 800 ms | 20-35Hz | 0.0028 | -1105.27 | 25.5–35.0 | 410–510 | 0.01 | T8C2z-T9B1z, T9A1z-T9C1z, T9B1z-T9C2z, T9C1z-T10A1z |
| CRNP-04 | Every Other | -400 to -100 ms | 0 to 800 ms | 20-35Hz | 0.005 | -860.72 | 20.0–27.5 | 630–800 | 0.01 | T8B1z-T8C2z, T8C1z-T9A1z, T8C2z-T9B1z, T9A1z-T9C1z |
| CRNP-04 | Every Other | -500 to -100 ms | 0 to 800 ms | 20-35Hz | 0.0002 | -3738.37 | 20.5–35.0 | 190–440 | 0.01 | T7C1z-T8B1z, T8A1z-T8C1z, T8B1z-T8C2z, T8C1z-T9A1z, T8C2z-T9B1z |
| CRNP-04 | Every Other | -500 to -100 ms | 0 to 800 ms | 20-35Hz | 0.0002 | -3731.14 | 20.0–35.0 | 0–170 | 0.01 | T7C1z-T8B1z, T8A1z-T8C1z, T8B1z-T8C2z, T8C1z-T9A1z, T8C2z-T9B1z, T9A1z-T9C1z |
| CRNP-04 | Every Other | -500 to -100 ms | 0 to 800 ms | 20-35Hz | 0.0002 | -2043.69 | 20.0–35.0 | 380–540 | 0.01 | T8A1z-T8C1z, T8B1z-T8C2z, T8C1z-T9A1z, T8C2z-T9B1z, T9A1z-T9C1z, T9B1z-T9C2z, T9C1z-T10A1z, T9C2z-T10B1z |
| CRNP-04 | Every Other | -500 to -100 ms | 0 to 800 ms | 20-35Hz | 0.0044 | -1063.09 | 20.0–32.5 | 620–800 | 0.01 | T8B1z-T8C2z, T8C1z-T9A1z, T8C2z-T9B1z, T9A1z-T9C1z |
| CRNP-04 | Every Other | -300 to -100 ms | 0 to 200 ms | 20-35Hz | 0.0002 | -1614.95 | 23.5–35.0 | 0–110 | 0.01 | T8A1z-T8C1z, T8B1z-T8C2z, T8C1z-T9A1z, T8C2z-T9B1z |
| CRNP-04 | Every Other | -400 to -100 ms | 0 to 200 ms | 20-35Hz | 0.0002 | -3197.32 | 20.0–35.0 | 0–180 | 0.01 | T7C1z-T8B1z, T8A1z-T8C1z, T8B1z-T8C2z, T8C1z-T9A1z, T8C2z-T9B1z, T9A1z-T9C1z, T9B1z-T9C2z |
| CRNP-04 | Every Other | -500 to -100 ms | 0 to 200 ms | 20-35Hz | 0.0002 | -3731.14 | 20.0–35.0 | 0–170 | 0.01 | T7C1z-T8B1z, T8A1z-T8C1z, T8B1z-T8C2z, T8C1z-T9A1z, T8C2z-T9B1z, T9A1z-T9C1z |
| CRNP-04 | Every Other | -500 to -100 ms | 0 to 200 ms | 20-35Hz | 0.0302 | -278.2 | 21.5–32.0 | 0–40 | 0.01 | T9B1z-T9C2z, T9C1z-T10A1z |
| CRNP-05 Lead 1 | Adjacent | -400 to -100 | 0 to 800 | 13-20Hz | 0.0028 | -1569.62 | 13.0-20.0 | 430-800 | 0.05 | T8A1L1-T8B1L1, T8B1L1-T8C1z, T8C1z-T9A1z, T9A1z-T9B1z, T9B1z-T9C1z, T9C1z-T9C3z |
| CRNP-05 Lead 1 | Adjacent | -400 to -100 | 0 to 800 | 13-20Hz | 0.0086 | -1164.24 | 13.0-20.0 | 0-410 | 0.05 | T8C1z-T9A1z, T9A1z-T9B1z, T9B1z-T9C1z, T9C1z-T9C3z |
| CRNP-05 Lead 1 | Adjacent | -500 to -100 | 0 to 800 | 13-20Hz | 0.001 | -2211.91 | 13.0-20.0 | 210-800 | 0.05 | T8A1L1-T8B1L1, T8B1L1-T8C1z, T8C1z-T9A1z, T9A1z-T9B1z, T9B1z-T9C1z, T9C1z-T9C3z |
| CRNP-05 Lead 1 | Adjacent | -500 to -100 | 0 to 800 | 13-20Hz | 0.004 | -1522.56 | 13.0-20.0 | 0-420 | 0.05 | T8C1z-T9A1z, T9A1z-T9B1z, T9B1z-T9C1z, T9C1z-T9C3z |
| CRNP-05 Lead 1 | Adjacent | -400 to -100 | 0 to 200 | 13-20Hz | 0.0324 | -415.94 | 13.5-20.0 | 0-200 | 0.05 | T8C1z-T9A1z, T9A1z-T9B1z, T9B1z-T9C1z |
| CRNP-05 Lead 1 | Adjacent | -500 to -100 | 0 to 200 | 13-20Hz | 0.0136 | -537.02 | 13.0-20.0 | 0-200 | 0.05 | T8C1z-T9A1z, T9A1z-T9B1z, T9B1z-T9C1z |
| CRNP-05 Lead 1 | Adjacent | -500 to -100 | 0 to 800 | 13-20Hz | 0.01 | -484.14 | 13.0-20.0 | 160-330 | 0.01 | T9A1z-T9B1z, T9B1z-T9C1z |
| CRNP-05 Lead 1 | Adjacent | -300 to -100 | 0 to 800 | 20-35Hz | 0.001 | -2833.28 | 21.5-35.0 | 0-250 | 0.05 | T9B1z-T9C1z, T9C1z-T9C3z, T9C3z-T10A2z |
| CRNP-05 Lead 1 | Adjacent | -300 to -100 | 0 to 800 | 20-35Hz | 0.0108 | -1589.89 | 22.0-35.0 | 440-680 | 0.05 | T8C1z-T9A1z, T9A1z-T9B1z, T9B1z-T9C1z, T9C1z-T9C3z, T9C3z-T10A2z |
| CRNP-05 Lead 1 | Adjacent | -400 to -100 | 0 to 800 | 20-35Hz | 0.0002 | -5003.88 | 21.0-35.0 | 0-260 | 0.05 | T8A1L1-T8B1L1, T8B1L1-T8C1z, T8C1z-T9A1z, T9A1z-T9B1z, T9B1z-T9C1z, T9C1z-T9C3z, T9C3z-T10A2z |
| CRNP-05 Lead 1 | Adjacent | -400 to -100 | 0 to 800 | 20-35Hz | 0.0196 | -1337.55 | 22.0-35.0 | 530-680 | 0.05 | T8B1L1-T8C1z, T8C1z-T9A1z, T9A1z-T9B1z, T9B1z-T9C1z, T9C1z-T9C3z, T9C3z-T10A2z |
| CRNP-05 Lead 1 | Adjacent | -400 to -100 | 0 to 800 | 20-35Hz | 0.0296 | -1160.2 | 21.0-35.0 | 290-490 | 0.05 | T8A1L1-T8B1L1, T8B1L1-T8C1z, T8C1z-T9A1z, T9A1z-T9B1z, T9B1z-T9C1z, T9C1z-T9C3z, T9C3z-T10A2z |
| CRNP-05 Lead 1 | Adjacent | -500 to -100 | 0 to 800 | 20-35Hz | 0.0002 | -6096.19 | 20.0-35.0 | 0-260 | 0.05 | T8A1L1-T8B1L1, T8B1L1-T8C1z, T8C1z-T9A1z, T9A1z-T9B1z, T9B1z-T9C1z, T9C1z-T9C3z, T9C3z-T10A2z |
| CRNP-05 Lead 1 | Adjacent | -500 to -100 | 0 to 800 | 20-35Hz | 0.0006 | -3892.69 | 20.0-35.0 | 290-680 | 0.05 | T8A1L1-T8B1L1, T8B1L1-T8C1z, T8C1z-T9A1z, T9A1z-T9B1z, T9B1z-T9C1z, T9C1z-T9C3z, T9C3z-T10A2z |
| CRNP-05 Lead 1 | Adjacent | -300 to -100 | 0 to 200 | 20-35Hz | 0.0002 | -2673.76 | 21.5-35.0 | 0-200 | 0.05 | T9B1z-T9C1z, T9C1z-T9C3z, T9C3z-T10A2z |
| CRNP-05 Lead 1 | Adjacent | -300 to -100 | 0 to 200 | 20-35Hz | 0.0142 | -862.88 | 23.5-35.0 | 10-200 | 0.05 | T8A1L1-T8B1L1, T8B1L1-T8C1z, T8C1z-T9A1z, T9A1z-T9B1z |
| CRNP-05 Lead 1 | Adjacent | -400 to -100 | 0 to 200 | 20-35Hz | 0.0002 | -4561.56 | 21.0-35.0 | 0-200 | 0.05 | T8A1L1-T8B1L1, T8B1L1-T8C1z, T8C1z-T9A1z, T9A1z-T9B1z, T9B1z-T9C1z, T9C1z-T9C3z, T9C3z-T10A2z |
| CRNP-05 Lead 1 | Adjacent | -500 to -100 | 0 to 200 | 20-35Hz | 0.0002 | -5591.91 | 20.0–35.0 | 0–200 | 0.05 | T8A1L1-T8B1L1, T8B1L1-T8C1z, T8C1z-T9A1z, T9A1z-T9B1z, T9B1z-T9C1z, T9C1z-T9C3z, T9C3z-T10A2z |
| CRNP-05 Lead 1 | Adjacent | -300 to -100 | 0 to 800 | 20-35Hz | 0.001 | -1357.02 | 24.5-35.0 | 10-230 | 0.01 | T9B1z-T9C1z, T9C1z-T9C3z, T9C3z-T10A2z |
| CRNP-05 Lead 1 | Adjacent | -400 to -100 | 0 to 800 | 20-35Hz | 0.0014 | -1038.38 | 24.5-35.0 | 20-230 | 0.01 | T9B1z-T9C1z, T9C1z-T9C3z, T9C3z-T10A2z |
| CRNP-05 Lead 1 | Adjacent | -400 to -100 | 0 to 800 | 20-35Hz | 0.002 | -967.24 | 22.0-35.0 | 20-210 | 0.01 | T8A1L1-T8B1L1, T8B1L1-T8C1z, T8C1z-T9A1z |
| CRNP-05 Lead 1 | Adjacent | -500 to -100 | 0 to 800 | 20-35Hz | 0.0004 | -1477.5 | 24.0-35.0 | 40-230 | 0.01 | T9B1z-T9C1z, T9C1z-T9C3z, T9C3z-T10A2z |
| CRNP-05 Lead 1 | Adjacent | -500 to -100 | 0 to 800 | 20-35Hz | 0.0004 | -1253.86 | 21.0-35.0 | 20-210 | 0.01 | T8A1L1-T8B1L1, T8B1L1-T8C1z, T8C1z-T9A1z |
| CRNP-05 Lead 1 | Adjacent | -300 to -100 | 0 to 200 | 20-35Hz | 0.0002 | -1293.76 | 24.5-35.0 | 10-200 | 0.01 | T9B1z-T9C1z, T9C1z-T9C3z, T9C3z-T10A2z |
| CRNP-05 Lead 1 | Adjacent | -400 to -100 | 0 to 200 | 20-35Hz | 0.0008 | -989.32 | 24.5-35.0 | 20-200 | 0.01 | T9B1z-T9C1z, T9C1z-T9C3z, T9C3z-T10A2z |
| CRNP-05 Lead 1 | Adjacent | -400 to -100 | 0 to 200 | 20-35Hz | 0.0008 | -928.01 | 22.0-35.0 | 20-200 | 0.01 | T8A1L1-T8B1L1, T8B1L1-T8C1z, T8C1z-T9A1z |
| CRNP-05 Lead 1 | Adjacent | -500 to -100 | 0 to 200 | 20-35Hz | 0.0002 | -1407.65 | 24.0-35.0 | 40-200 | 0.01 | T9B1z-T9C1z, T9C1z-T9C3z, T9C3z-T10A2z |
| CRNP-05 Lead 1 | Adjacent | -500 to -100 | 0 to 200 | 20-35Hz | 0.0002 | -1218.43 | 21.0-35.0 | 20-200 | 0.01 | T8A1L1-T8B1L1, T8B1L1-T8C1z, T8C1z-T9A1z |
| CRNP-05 Lead 1 | Every Other | -400 to -100 ms | 0 to 800 ms | 13-20Hz | 0.0022 | -2520.21 | 13.0–20.0 | 50–800 | 0.05 | T8A1L1-T8C1z, T8B1L1-T9A1z, T8C1z-T9B1z, T9A1z-T9C1z |
| CRNP-05 Lead 1 | Every Other | -500 to -100 ms | 0 to 800 ms | 13-20Hz | 0.0004 | -3260.73 | 13.0–20.0 | 40–800 | 0.05 | T8A1L1-T8C1z, T8B1L1-T9A1z, T8C1z-T9B1z, T9A1z-T9C1z, T9B1z-T9C3z, T9C1z-T10A2z |
| CRNP-05 Lead 1 | Every Other | -400 to -100 ms | 0 to 200 ms | 13-20Hz | 0.0264 | -533.58 | 13.0–20.0 | 50–200 | 0.05 | T8B1L1-T9A1z, T8C1z-T9B1z, T9A1z-T9C1z |
| CRNP-05 Lead 1 | Every Other | -500 to -100 ms | 0 to 200 ms | 13-20Hz | 0.0178 | -644.72 | 13.0–20.0 | 40–200 | 0.05 | T8B1L1-T9A1z, T8C1z-T9B1z, T9A1z-T9C1z, T9B1z-T9C3z |
| CRNP-05 Lead 1 | Every Other | -500 to -100 ms | 0 to 200 ms | 13-20Hz | 0.022 | -585.45 | 13.0–20.0 | 0–200 | 0.05 | T8A1L1-T8C1z, T8B1L1-T9A1z |
| CRNP-05 Lead 1 | Every Other | -300 to -100 ms | 0 to 800 ms | 20-35Hz | 0.0004 | -3464.23 | 20.0–35.0 | 0–220 | 0.05 | T8A1L1-T8C1z, T8B1L1-T9A1z, T8C1z-T9B1z, T9A1z-T9C1z, T9B1z-T9C3z, T9C1z-T10A2z |

| Subject | Bipolar montage | Baseline | Post-stim | Beta band | P Value | Cluster Stat | Cluster freq (Hz) | Time (ms) | Cluster-α | Channels |
|---|---|---|---|---|---|---|---|---|---|---|
| CRNP-05 Lead 1 | Every Other | -400 to -100 ms | 0 to 800 ms | 20-35Hz | 0.0002 | -5700.71 | 20.0–35.0 | 0–270 | 0.05 | T8A1L1-T8C1z, T8B1L1-T9A1z, T8C1z-T9B1z, T9A1z-T9C1z, T9B1z-T9C3z, T9C1z-T10A2z |
| CRNP-05 Lead 1 | Every Other | -500 to -100 ms | 0 to 800 ms | 20-35Hz | 0.0004 | -6433.54 | 20.0–35.0 | 0–260 | 0.05 | T8A1L1-T8C1z, T8B1L1-T9A1z, T8C1z-T9B1z, T9A1z-T9C1z, T9B1z-T9C3z, T9C1z-T10A2z |
| CRNP-05 Lead 1 | Every Other | -300 to -100 ms | 0 to 200 ms | 20-35Hz | 0.0002 | -3397.82 | 20.0–35.0 | 0–200 | 0.05 | T8A1L1-T8C1z, T8B1L1-T9A1z, T8C1z-T9B1z, T9A1z-T9C1z, T9B1z-T9C3z, T9C1z-T10A2z |
| CRNP-05 Lead 1 | Every Other | -400 to -100 ms | 0 to 200 ms | 20-35Hz | 0.0002 | -5283.4 | 20.0–35.0 | 0–200 | 0.05 | T8A1L1-T8C1z, T8B1L1-T9A1z, T8C1z-T9B1z, T9A1z-T9C1z, T9B1z-T9C3z, T9C1z-T10A2z |
| CRNP-05 Lead 1 | Every Other | -500 to -100 ms | 0 to 200 ms | 20-35Hz | 0.0002 | -6102.27 | 20.0–35.0 | 0–200 | 0.05 | T8A1L1-T8C1z, T8B1L1-T9A1z, T8C1z-T9B1z, T9A1z-T9C1z, T9B1z-T9C3z, T9C1z-T10A2z |
| CRNP-05 Lead 1 | Every Other | -300 to -100 ms | 0 to 800 ms | 20-35Hz | 0.0236 | -479.16 | 21.5–33.0 | 20–130 | 0.01 | T8A1L1-T8C1z, T8B1L1-T9A1z, T8C1z-T9B1z |
| CRNP-05 Lead 1 | Every Other | -300 to -100 ms | 0 to 800 ms | 20-35Hz | 0.0318 | -425.91 | 24.5–32.0 | 0–110 | 0.01 | T9A1z-T9C1z, T9B1z-T9C3z |
| CRNP-05 Lead 1 | Every Other | -400 to -100 ms | 0 to 800 ms | 20-35Hz | 0.0004 | -1459.83 | 20.0–35.0 | 0–150 | 0.01 | T8A1L1-T8C1z, T8B1L1-T9A1z, T8C1z-T9B1z |
| CRNP-05 Lead 1 | Every Other | -400 to -100 ms | 0 to 800 ms | 20-35Hz | 0.0446 | -388.49 | 23.5–31.0 | 0–90 | 0.01 | T9A1z-T9C1z, T9B1z-T9C3z |
| CRNP-05 Lead 1 | Every Other | -500 to -100 ms | 0 to 800 ms | 20-35Hz | 0.0006 | -1948.97 | 20.0–35.0 | 0–160 | 0.01 | T8A1L1-T8C1z, T8B1L1-T9A1z, T8C1z-T9B1z |
| CRNP-05 Lead 1 | Every Other | -500 to -100 ms | 0 to 800 ms | 20-35Hz | 0.0204 | -513.33 | 24.0–32.5 | 10–110 | 0.01 | T9A1z-T9C1z, T9B1z-T9C3z |
| CRNP-05 Lead 1 | Every Other | -300 to -100 ms | 0 to 200 ms | 20-35Hz | 0.0066 | -479.16 | 21.5–33.0 | 20–130 | 0.01 | T8A1L1-T8C1z, T8B1L1-T9A1z, T8C1z-T9B1z |
| CRNP-05 Lead 1 | Every Other | -300 to -100 ms | 0 to 200 ms | 20-35Hz | 0.0102 | -425.91 | 24.5–32.0 | 0–110 | 0.01 | T9A1z-T9C1z, T9B1z-T9C3z |
| CRNP-05 Lead 1 | Every Other | -300 to -100 ms | 0 to 200 ms | 20-35Hz | 0.0198 | -311.71 | 27.5–35.0 | 140–200 | 0.01 | T9A1z-T9C1z, T9B1z-T9C3z, T9C1z-T10A2z |
| CRNP-05 Lead 1 | Every Other | -400 to -100 ms | 0 to 200 ms | 20-35Hz | 0.0002 | -1459.83 | 20.0–35.0 | 0–150 | 0.01 | T8A1L1-T8C1z, T8B1L1-T9A1z, T8C1z-T9B1z |
| CRNP-05 Lead 1 | Every Other | -400 to -100 ms | 0 to 200 ms | 20-35Hz | 0.0118 | -388.49 | 23.5–31.0 | 0–90 | 0.01 | T9A1z-T9C1z, T9B1z-T9C3z |
| CRNP-05 Lead 1 | Every Other | -400 to -100 ms | 0 to 200 ms | 20-35Hz | 0.0256 | -276.36 | 27.5–35.0 | 140–200 | 0.01 | T9A1z-T9C1z, T9B1z-T9C3z, T9C1z-T10A2z |
| CRNP-05 Lead 1 | Every Other | -500 to -100 ms | 0 to 200 ms | 20-35Hz | 0.0002 | -1948.97 | 20.0–35.0 | 0–160 | 0.01 | T8A1L1-T8C1z, T8B1L1-T9A1z, T8C1z-T9B1z |
| CRNP-05 Lead 1 | Every Other | -500 to -100 ms | 0 to 200 ms | 20-35Hz | 0.007 | -513.33 | 24.0–32.5 | 10–110 | 0.01 | T9A1z-T9C1z, T9B1z-T9C3z |
| CRNP-05 Lead 1 | Every Other | -500 to -100 ms | 0 to 200 ms | 20-35Hz | 0.02 | -337.7 | 27.0–35.0 | 140–200 | 0.01 | T9A1z-T9C1z, T9B1z-T9C3z, T9C1z-T10A2z |
| CRNP-05 Lead 2 | Adjacent | -400 to -100 ms | 0 to 800 ms | 13-20Hz | 0.0006 | -1888.4 | 13.0–20.0 | 0–450 | 0.05 | T9A2z-T9B2z, T9B2z-T9C2z |
| CRNP-05 Lead 2 | Adjacent | -500 to -100 ms | 0 to 800 ms | 13-20Hz | 0.0008 | -2115.73 | 13.0–20.0 | 0–460 | 0.05 | T9A2z-T9B2z, T9B2z-T9C2z, T9C2z-T10A1z |
| CRNP-05 Lead 2 | Adjacent | -500 to -100 ms | 0 to 800 ms | 13-20Hz | 0.0018 | -1431.34 | 13.0–20.0 | 510–800 | 0.05 | T9A2z-T9B2z, T9B2z-T9C2z |
| CRNP-05 Lead 2 | Adjacent | -400 to -100 ms | 0 to 200 ms | 13-20Hz | 0.0018 | -927.42 | 13.0–20.0 | 0–200 | 0.05 | T9A2z-T9B2z, T9B2z-T9C2z |
| CRNP-05 Lead 2 | Adjacent | -500 to -100 ms | 0 to 200 ms | 13-20Hz | 0.001 | -956.58 | 13.0–20.0 | 0–200 | 0.05 | T9A2z-T9B2z, T9B2z-T9C2z |
| CRNP-05 Lead 2 | Adjacent | -300 to -100 ms | 0 to 800 ms | 20-35Hz | 0.0002 | -11209.05 | 20.5–35.0 | 0–580 | 0.05 | T9A2z-T9B2z, T9B2z-T9C2z, T9C2z-T10A1z, T10A1z-T10A3z, T10A3z-T10B1z, T10B1z-T10C1z, T10C1z-T11A1z |
| CRNP-05 Lead 2 | Adjacent | -300 to -100 ms | 0 to 800 ms | 20-35Hz | 0.0256 | -1087.34 | 20.0–35.0 | 580–800 | 0.05 | T10A1z-T10A3z, T10A3z-T10B1z, T10B1z-T10C1z, T10C1z-T11A1z |
| CRNP-05 Lead 2 | Adjacent | -400 to -100 ms | 0 to 800 ms | 20-35Hz | 0.0002 | -11764.45 | 20.0–35.0 | 0–570 | 0.05 | T9A2z-T9B2z, T9B2z-T9C2z, T9C2z-T10A1z, T10A1z-T10A3z, T10A3z-T10B1z, T10B1z-T10C1z, T10C1z-T11A1z |
| CRNP-05 Lead 2 | Adjacent | -400 to -100 ms | 0 to 800 ms | 20-35Hz | 0.0078 | -1854.76 | 20.0–35.0 | 530–800 | 0.05 | T9A2z-T9B2z, T9B2z-T9C2z, T9C2z-T10A1z, T10A1z-T10A3z, T10A3z-T10B1z |
| CRNP-05 Lead 2 | Adjacent | -400 to -100 ms | 0 to 800 ms | 20-35Hz | 0.028 | -1117.52 | 20.0–35.0 | 590–800 | 0.05 | T10A3z-T10B1z, T10B1z-T10C1z, T10C1z-T11A1z |
| CRNP-05 Lead 2 | Adjacent | -500 to -100 ms | 0 to 800 ms | 20-35Hz | 0.0002 | -12474.55 | 20.0–35.0 | 0–560 | 0.05 | T9A2z-T9B2z, T9B2z-T9C2z, T9C2z-T10A1z, T10A1z-T10A3z, T10A3z-T10B1z, T10B1z-T10C1z, T10C1z-T11A1z |
| CRNP-05 Lead 2 | Adjacent | -500 to -100 ms | 0 to 800 ms | 20-35Hz | 0.0018 | -3447.81 | 20.0–35.0 | 500–800 | 0.05 | T9A2z-T9B2z, T9B2z-T9C2z, T9C2z-T10A1z, T10A1z-T10A3z, T10A3z-T10B1z, T10B1z-T10C1z, T10C1z-T11A1z |
| CRNP-05 Lead 2 | Adjacent | -300 to -100 ms | 0 to 200 ms | 20-35Hz | 0.0002 | -6603.41 | 20.5–35.0 | 0–200 | 0.05 | T9A2z-T9B2z, T9B2z-T9C2z, T9C2z-T10A1z, T10A1z-T10A3z, T10A3z-T10B1z, T10B1z-T10C1z, T10C1z-T11A1z |
| CRNP-05 Lead 2 | Adjacent | -400 to -100 ms | 0 to 200 ms | 20-35Hz | 0.0002 | -7076.81 | 20.0–35.0 | 0–200 | 0.05 | T9A2z-T9B2z, T9B2z-T9C2z, T9C2z-T10A1z, T10A1z-T10A3z, T10A3z-T10B1z, T10B1z-T10C1z, T10C1z-T11A1z |
| CRNP-05 Lead 2 | Adjacent | -500 to -100 ms | 0 to 200 ms | 20-35Hz | 0.0002 | -7552.14 | 20.0–35.0 | 0–200 | 0.05 | T9A2z-T9B2z, T9B2z-T9C2z, T9C2z-T10A1z, T10A1z-T10A3z, T10A3z-T10B1z, T10B1z-T10C1z, T10C1z-T11A1z |
| CRNP-05 Lead 2 | Adjacent | -300 to -100 ms | 0 to 800 ms | 20-35Hz | 0.0002 | -3572.96 | 23.0–35.0 | 0–220 | 0.01 | T9A2z-T9B2z, T9B2z-T9C2z, T9C2z-T10A1z, T10A1z-T10A3z, T10A3z-T10B1z, T10B1z-T10C1z, T10C1z-T11A1z |
| CRNP-05 Lead 2 | Adjacent | -300 to -100 ms | 0 to 800 ms | 20-35Hz | 0.0002 | -1452.63 | 23.5–35.0 | 290–490 | 0.01 | T9B2z-T9C2z, T9C2z-T10A1z, T10A1z-T10A3z, T10A3z-T10B1z, T10B1z-T10C1z |
| CRNP-05 Lead 2 | Adjacent | -400 to -100 ms | 0 to 800 ms | 20-35Hz | 0.0002 | -3800.66 | 22.5–35.0 | 0–240 | 0.01 | T9A2z-T9B2z, T9B2z-T9C2z, T9C2z-T10A1z, T10A1z-T10A3z, T10A3z-T10B1z, T10B1z-T10C1z, T10C1z-T11A1z |
| CRNP-05 Lead 2 | Adjacent | -400 to -100 ms | 0 to 800 ms | 20-35Hz | 0.008 | -551.99 | 24.0–35.0 | 380–490 | 0.01 | T9B2z-T9C2z, T9C2z-T10A1z |
| CRNP-05 Lead 2 | Adjacent | -400 to -100 ms | 0 to 800 ms | 20-35Hz | 0.027 | -379.64 | 25.0–32.5 | 350–470 | 0.01 | T9C2z-T10A1z, T10A1z-T10A3z, T10A3z-T10B1z, T10B1z-T10C1z |
| CRNP-05 Lead 2 | Adjacent | -500 to -100 ms | 0 to 800 ms | 20-35Hz | 0.0002 | -3795.01 | 22.0–35.0 | 0–240 | 0.01 | T9C2z-T10A1z, T10A1z-T10A3z, T10A3z-T10B1z, T10B1z-T10C1z, T10C1z-T11A1z |
| CRNP-05 Lead 2 | Adjacent | -500 to -100 ms | 0 to 800 ms | 20-35Hz | 0.0004 | -1056.23 | 24.0–35.0 | 320–480 | 0.01 | T9B2z-T9C2z, T9C2z-T10A1z, T10A1z-T10A3z, T10A3z-T10B1z, T10B1z-T10C1z |
| CRNP-05 Lead 2 | Adjacent | -500 to -100 ms | 0 to 800 ms | 20-35Hz | 0.0038 | -611.75 | 21.5–34.5 | 70–220 | 0.01 | T9A2z-T9B2z, T9B2z-T9C2z |
| CRNP-05 Lead 2 | Adjacent | -300 to -100 ms | 0 to 200 ms | 20-35Hz | 0.0002 | -3510.86 | 23.0–35.0 | 0–200 | 0.01 | T9A2z-T9B2z, T9B2z-T9C2z, T9C2z-T10A1z, T10A1z-T10A3z, T10A3z-T10B1z, T10B1z-T10C1z, T10C1z-T11A1z |
| CRNP-05 Lead 2 | Adjacent | -400 to -100 ms | 0 to 200 ms | 20-35Hz | 0.0002 | -3693.77 | 22.5–35.0 | 0–200 | 0.01 | T9A2z-T9B2z, T9B2z-T9C2z, T9C2z-T10A1z, T10A1z-T10A3z, T10A3z-T10B1z, T10B1z-T10C1z, T10C1z-T11A1z |
| CRNP-05 Lead 2 | Adjacent | -500 to -100 ms | 0 to 200 ms | 20-35Hz | 0.0002 | -3641.49 | 22.0–35.0 | 0–200 | 0.01 | T9C2z-T10A1z, T10A1z-T10A3z, T10A3z-T10B1z, T10B1z-T10C1z, T10C1z-T11A1z |
| CRNP-05 Lead 2 | Adjacent | -500 to -100 ms | 0 to 200 ms | 20-35Hz | 0.0036 | -561.61 | 21.5–34.5 | 70–200 | 0.01 | T9A2z-T9B2z, T9B2z-T9C2z |
| CRNP-05 Lead 2 | Every Other | -500 to -100 ms | 0 to 800 ms | 13-20Hz | 0.007 | -1213.28 | 13.0–19.0 | 70–800 | 0.05 | T9C2z-T10A3z, T10A1z-T10B1z, T10A3z-T10C1z, T10B1z-T11A1z |
| CRNP-05 Lead 2 | Every Other | -500 to -100 ms | 0 to 800 ms | 13-20Hz | 0.015 | -999.21 | 13.0–20.0 | 110–460 | 0.05 | T9A2z-T9C2z, T9B2z-T10A1z |
| CRNP-05 Lead 2 | Every Other | -500 to -100 ms | 0 to 200 ms | 13-20Hz | 0.0382 | -374.28 | 13.0–20.0 | 110–200 | 0.05 | T9A2z-T9C2z, T9B2z-T10A1z |
| CRNP-05 Lead 2 | Every Other | -500 to -100 ms | 0 to 200 ms | 13-20Hz | 0.0382 | -373.95 | 13.0–19.0 | 70–200 | 0.05 | T9C2z-T10A3z, T10A1z-T10B1z, T10A3z-T10C1z, T10B1z-T11A1z |
| CRNP-05 Lead 2 | Every Other | -300 to -100 ms | 0 to 800 ms | 20-35Hz | 0.0002 | -6230.14 | 22.0–35.0 | 0–230 | 0.05 | T9A2z-T9C2z, T9B2z-T10A1z, T9C2z-T10A3z, T10A1z-T10B1z, T10A3z-T10C1z, T10B1z-T11A1z |
| CRNP-05 Lead 2 | Every Other | -300 to -100 ms | 0 to 800 ms | 20-35Hz | 0.016 | -1860.47 | 20.0–35.0 | 260–490 | 0.05 | T9A2z-T9C2z, T9B2z-T10A1z, T9C2z-T10A3z, T10A1z-T10B1z, T10A3z-T10C1z, T10B1z-T11A1z |
| CRNP-05 Lead 2 | Every Other | -400 to -100 ms | 0 to 800 ms | 20-35Hz | 0.0002 | -6501.49 | 20.0–35.0 | 0–240 | 0.05 | T9A2z-T9C2z, T9B2z-T10A1z, T9C2z-T10A3z, T10A1z-T10B1z, T10A3z-T10C1z, T10B1z-T11A1z |
| CRNP-05 Lead 2 | Every Other | -400 to -100 ms | 0 to 800 ms | 20-35Hz | 0.0238 | -1692.75 | 20.0–35.0 | 320–510 | 0.05 | T9A2z-T9C2z, T9B2z-T10A1z, T9C2z-T10A3z, T10A1z-T10B1z, T10A3z-T10C1z, T10B1z-T11A1z |
| CRNP-05 Lead 2 | Every Other | -500 to -100 ms | 0 to 800 ms | 20-35Hz | 0.0002 | -6767.1 | 20.0–35.0 | 0–240 | 0.05 | T9A2z-T9C2z, T9B2z-T10A1z, T9C2z-T10A3z, T10A1z-T10B1z, T10A3z-T10C1z, T10B1z-T11A1z |

| Subject | Bipolar montage | Baseline | Post-stim | Beta band | P Value | Cluster Stat | Cluster freq (Hz) | Time (ms) | Cluster-α | Channels |
|---|---|---|---|---|---|---|---|---|---|---|
| CRNP-05 Lead 2 | Every Other | -500 to -100 ms | 0 to 800 ms | 20-35Hz | 0.0286 | -1594.78 | 20.0–35.0 | 330–480 | 0.05 | T9A2z-T9C2z, T9B2z-T10A1z, T9C2z-T10A3z, T10A1z-T10B1z, T10A3z-T10C1z, T10B1z-T11A1z |
| CRNP-05 Lead 2 | Every Other | -300 to -100 ms | 0 to 200 ms | 20-35Hz | 0.0002 | -5973.57 | 22.0–35.0 | 0–200 | 0.05 | T9A2z-T9C2z, T9B2z-T10A1z, T9C2z-T10A3z, T10A1z-T10B1z, T10A3z-T10C1z, T10B1z-T11A1z |
| CRNP-05 Lead 2 | Every Other | -400 to -100 ms | 0 to 200 ms | 20-35Hz | 0.0002 | -6204.09 | 20.0–35.0 | 0–200 | 0.05 | T9A2z-T9C2z, T9B2z-T10A1z, T9C2z-T10A3z, T10A1z-T10B1z, T10A3z-T10C1z, T10B1z-T11A1z |
| CRNP-05 Lead 2 | Every Other | -500 to -100 ms | 0 to 200 ms | 20-35Hz | 0.0002 | -6501.95 | 20.0–35.0 | 0–200 | 0.05 | T9A2z-T9C2z, T9B2z-T10A1z, T9C2z-T10A3z, T10A1z-T10B1z, T10A3z-T10C1z, T10B1z-T11A1z |
| CRNP-05 Lead 2 | Every Other | -300 to -100 ms | 0 to 800 ms | 20-35Hz | 0.0004 | -3446.56 | 22.5–35.0 | 0–210 | 0.01 | T9A2z-T9C2z, T9B2z-T10A1z, T9C2z-T10A3z, T10A1z-T10B1z, T10A3z-T10C1z, T10B1z-T11A1z |
| CRNP-05 Lead 2 | Every Other | -300 to -100 ms | 0 to 800 ms | 20-35Hz | 0.046 | -361.92 | 23.0–35.0 | 400–470 | 0.01 | T9A2z-T9C2z, T9B2z-T10A1z |
| CRNP-05 Lead 2 | Every Other | -400 to -100 ms | 0 to 800 ms | 20-35Hz | 0.0002 | -3620.49 | 23.0–35.0 | 0–210 | 0.01 | T9A2z-T9C2z, T9B2z-T10A1z, T9C2z-T10A3z, T10A1z-T10B1z, T10A3z-T10C1z, T10B1z-T11A1z |
| CRNP-05 Lead 2 | Every Other | -500 to -100 ms | 0 to 800 ms | 20-35Hz | 0.0002 | -3562.97 | 23.0–35.0 | 0–210 | 0.01 | T9A2z-T9C2z, T9B2z-T10A1z, T9C2z-T10A3z, T10A1z-T10B1z, T10A3z-T10C1z, T10B1z-T11A1z |
| CRNP-05 Lead 2 | Every Other | -300 to -100 ms | 0 to 200 ms | 20-35Hz | 0.0002 | -3417.6 | 22.5–35.0 | 0–200 | 0.01 | T9A2z-T9C2z, T9B2z-T10A1z, T9C2z-T10A3z, T10A1z-T10B1z, T10A3z-T10C1z, T10B1z-T11A1z |
| CRNP-05 Lead 2 | Every Other | -400 to -100 ms | 0 to 200 ms | 20-35Hz | 0.0002 | -3588.92 | 23.0–35.0 | 0–200 | 0.01 | T9A2z-T9C2z, T9B2z-T10A1z, T9C2z-T10A3z, T10A1z-T10B1z, T10A3z-T10C1z, T10B1z-T11A1z |
| CRNP-05 Lead 2 | Every Other | -500 to -100 ms | 0 to 200 ms | 20-35Hz | 0.0002 | -3534.33 | 23.0–35.0 | 0–200 | 0.01 | T9A2z-T9C2z, T9B2z-T10A1z, T9C2z-T10A3z, T10A1z-T10B1z, T10A3z-T10C1z, T10B1z-T11A1z |

**Supplementary Table 1. Summary table of statistically significant low- and high-beta suppression clusters for adjacent and every-other contact bipolar channels for a range of baseline reference and post-stimulus windows for all subjects/leads.** Results are shown at cluster forming thresholds (Cluster-α) of either $p < 0.05$ or $p < 0.01$.
